# Low-dose chemotherapy combined with delayed immunotherapy in the neoadjuvant treatment of non-small cell lung cancer and dynamic monitoring of the drug response in peripheral blood

**DOI:** 10.1101/2024.07.11.24310105

**Authors:** Chaoyang Liang, Qi Song, Wenhao Zhou, Na Li, Qi Xiong, Chaohu Pan, Shaohong Zhao, Xiang Yan, Xiaoling Zhang, Yaping Long, Juntang Guo, Tao Wang, Weiwei Shi, Shengjie Sun, Bo Yang, Zhouhuan Dong, Haitao Luo, Jie Li, Yi Hu, Bo Yang

## Abstract

**Background:** Despite chemo-immunotherapy has been applied to the neoadjuvant treatment of non-small cell lung cancer (NSCLC), the impacts of dosage and the order of medication on treatment efficacy and safety remain largely unexplored. We originally designed an exploratory study to investigate the efficacy and safety of reduced-dose chemotherapy combined with delayed immunotherapy as well as the dynamic changes of circulating tumor DNA (ctDNA) and T cell receptor (TCR) during the therapy.

**Methods:** Patients with clinical stage IIA to IIIA resectable NSCLC were treated with 2 cycles of reduced-dose platinum-based chemotherapy on day 1 combined with immunotherapy on day 5. The same postoperative modified adjuvant therapy regimen was administered for 2 cycles. Plasma samples at different time-points were collected and performed with T cell receptor (TCR) and circulating tumor DNA (ctDNA) sequencing.

**Results:** 38 patients received modified chemo-immunotherapy. The proportion of patients exhibiting complete response and partial response was 5.3% and 68.4%, respectively. The confirmed objective response rate was 73.7%. Radiological downstaging was achieved in 39.5%. Major pathologic response and complete pathologic response were observed in 47.4% and 31.6% of patients, respectively. Only one patient experienced grade 3 adverse event. Further analyses revealed that this modified chemo-immunotherapy led to the expansion of predominant TCR clones and reduction of tumor burden after the first cycle of chemotherapy.

**Conclusion:** The promising clinical efficacy and low side effects of modified neoadjuvant chemo-immunotherapy position it as a prospective and innovative strategy for NSCLC.

**Trial registration:** Registration Number: ChiCTR2000033092

## Background

Approximately 20 to 25% of patients diagnosed with NSCLC are eligible for surgical resection (1), yet 30% to 55% of those who undergo curative surgery will encounter recurrence and ultimately die (2, 3). Despite the improvement in treatment of NSCLC patients, the 5-year survival rates remain relatively low, hovering around 36% (4). Over the past decade, neoadjuvant platinum-based chemotherapy, followed by surgery, has become the standard of care. Nevertheless, only 5 to 6 % improvements have been noted in 5-year recurrence-free survival and overall survival rates compared to surgery alone (5).

Currently, combination of chemotherapy with immunotherapy has demonstrated superior clinical efficacy and prolonged event-free survival for patients with resectable NSCLC. Trials such as CheckMate 816 (6), SAKK 16/14 (7), and NADIM (8), have showcased the enhanced efficacy and manageable safety of combination therapy compared to monotherapy. However, for most previous clinical trials, cytotoxic drugs with standard doses and anti-PD1 antibody were administrated on the same day (9), which may result in unsatisfactory eradication rates and relatively high incidence of severe treatment-related adverse events (TRAEs). To improve the cure rate and reduce TRAEs, the use of low-dose chemotherapy may represent a feasible alternative, given it favorable tolerability profile without compromising efficacy (10, 11). Although various regimens of low-dose chemotherapy have been utilized in cancer therapy (12, 13), the effects of dosage and administration sequence on the efficacy and safety of NSCLC treatment remain controversial. According to our previous research findings, the administration of immunotherapy 3-5 days after chemotherapy significantly reduces lymphocyte damage (14). Therefore, we hypothesize that by coordinating the dosage of chemotherapy and the timing of immunotherapy administration, we can enhance the synergistic effects between the two interventions. Ultimately, this could improve clinical efficacy while reducing toxic side effects.

To monitor the therapeutic efficacy during the process of chemo-immunotherapy, there is an urgent requirement for reliable indicators. The T cell receptor (TCR) repertoire and the presence of circulating tumor DNA (ctDNA) in blood have been proven to be efficient predictors of therapeutic responses in cancer treatment (15, 16). Chemotherapeutic agents have the capability to induce immunogenic death of tumor cells, reduce tumor burden, and promote the infiltration of CD8 T lymphocyte (17, 18). The recognition of tumor antigens by CD8 T lymphocytes relies primarily on the presence of a diverse and polymorphic TCR within an individual (19). Hence, it is imperative to explore the dynamic changes of TCR diversity and tumor burden in ctDNA throughout the course of therapy.

In this study, we report the results of our pilot clinical trial assessing the efficacy and safety of modified neoadjuvant chemo-immunotherapy in patients with resectable NSCLC. These results indicate that the combination of low-dose chemotherapy followed by delayed immunotherapy could potentially induce tumor cell damage, promote antigen exposure, and support T cell expansion, thereby enhancing therapeutic efficacy and reducing toxic side effects. Monitoring the dynamic changes of TCR repertoire and ctDNA does not only offer insights into the mechanism underlying our therapeutic approach to some extent but also furnishes valuable data for monitoring treatment efficacy.

## Materials and Methods

### Study design and participants

38 Patients with resectable stage IIA to IIIA NSCLC, as per the staging criteria outlined by the American Joint Committee on Cancer, 8th edition, and possessing adequate organ function, along with an Eastern Cooperative Oncology Group performance-status rating of 0 to 1, were included in this study. All patients had not received any prior cancer treatment, exhibited measurable disease according to the Response Evaluation Criteria in Solid Tumors (version 1.1) and had confirmed histological or cytological evidence of the disease. Exclusion criteria included the presence of identified *EGFR* mutations or *ALK* translocations; a diagnosis of malignancies beyond NSCLC within 5 years; previous exposure to specific therapies such as anti-PD-1, anti-PD-L1, or anti-PD-L2 agents or medications targeting T cell receptor modulation (e.g., CTLA-4, LAG-3, or TIM-3); a history of allogeneic organ or hematopoietic stem cell transplantation (except corneal transplantation); ongoing repercussions from prior interventions; pregnancy or breastfeeding; and HIV positivity. Moreover, the availability of pre- and post-adjuvant treatment tumor tissues and serial blood samples facilitated comprehensive analyses including whole transcriptome sequencing, TCR sequencing, and ctDNA sequencing.

### Trial design and treatment

This study was a single-center, single-arm, pilot clinical trial (Registration Number: ChiCTR2000033092). Eligible patients received 2 cycles of neoadjuvant therapy of low-dose platinum-based chemotherapy: albumin-bound paclitaxel 180-220 mg/m^2^ plus cisplatin 60 mg/m^2^ for lung squamous cell carcinoma or pemetrexed disodium 500 mg/m^2^ plus cisplatin 60 mg/m^2^ for lung adenocarcinoma respectively on day 1 plus sintilimab 200 mg on day 5 of each cycle, followed by radiographic evaluation before undergoing radical surgery. Surgery was conducted within 6 weeks after the completion of neoadjuvant treatment. Then patients would receive 2 cycles of adjuvant therapy with the same regimen.

### Study outcomes

The primary endpoints include objective response rate (ORR: the proportion of patients with complete response (CR) or partial response (PR) according to response evaluation criteria in solid tumors) and major pathologic response (MPR) (less than 10% residual viable tumor cells in the primary tumor and sampled lymph nodes) according to the pathologist review. Secondary endpoint included pathologic complete response (pCR) (0% residual viable tumor cells in the primary tumor and sampled lymph nodes), disease free survival (DFS), treatment-related adverse events (TRAEs), occurrence of surgical complications, and intraoperative and perioperative complications.

### Plasma collection, cell-free DNA extraction, and sequencing

Blood samples were collected into cell-free DNA blood streck tubes. Isolated plasma and paired white blood cell (WBC) control were stored at -80 until extraction. Cell-free DNA (cfDNA) and WBC-derived DNA were extracted from 4 mL plasma and WBC using QIAamp Circulating Nucleic Acid Kit (Qiagen, Hilden, Germany) and Mag-Bind® Blood & Tissue DNA HDQ 96 kit (OMEGA) according to the manufacturer’s instructions, respectively. DNA extracts were sheared to 100 bp fragments using an S220 focused ultrasonicator (Covaris, Woodbrun, MA, USA). The adaptors (NadPrep®) were attached to the terminal of fragments and followed by PCR (Bio-Rad T100, USA) replication. The libraries were constructed using NadPrep Plasma Cell-Free DNA Dual-Ended Molecular Tag Library Construction Kit (for MGI) (Nanodigmbio). The target customized probes YuceOne Plus (ctDNA) synthesized by IDT were used for hybridization enrichment. Target enrichment was performed with NadPrep Hybrid Capture Reagents (Nanodigmbio). The enriched DNA fragments were sequenced with the MGISEQ platform. The mean depth of coverage was 3445× for cfDNA and 1419× for matched peripheral blood samples.

### Circulating tumor DNA (ctDNA) analysis

SOAPnuke was implemented to cut the adapter and remove low-quality raw reads (20). Genecore was performed for cluster reads by unique molecule index (UMI) and remove the replicated reads derived from PCR (21). Clean reads were aligned against the human reference genome (hg19) with BWA (v0.7.12) (22) and duplicated reads were removed by samtools (v0.1.19) (23). Subsequently, generated BAM files were used for downstream analysis. We compared the ctDNA samples and matched control blood sequencing data to identify the somatic mutations, including single nucleotide variants (SNVs) and small insertions and deletions (Indels) using mutation caller Vardict (v1.7.0) with default parameters (24). Next, the caller was run with dbSNP (version 147), 1000G (phase3_release_v5), CLINVAR (version 151 ClinVar (nih.gov)), and COSMIC (version 81) data for known polymorphic sites. Substitutions and indels would be filtered out according to the following criteria: 1) low variant allelic fractions (VAF < 0.0005), 2) removed any variants with greater than 1 supporting read in the matched germline sample, 3) variants present in the above genome database. In addition, the filtered mutations were annotated by snpEff (v4.3) (25) with NCBIrefseq (https://www.ncbi.nlm.nih.gov/refseq/). The maximum VAF is considered as the tumor burden during the therapy.

### Whole transcriptome sequencing

The total RNA of tumor samples was isolated using the RNeasy FFPE Kit (Qiagen, GER) according to the manufacturer’s instructions. The concentrations of RNA were quantified using Qubit™ RNA HS Assay Kit (ThermoFisher Scientific, USA). RNA purity and integrity were checked using the Take3 (BioTek, USA) and the RNA Cartridge kit of the Qseq100 Bio-Fragment Analyzer (Bioptic, CHN), respectively. Then, RNA sequencing (RNA-seq) libraries were constructed using the rRNA depletion module (H/M/R) and NadPrep DNA Library Preparation Module for MGI (MGI, CHN). The libraries were sequenced on the MGISEQ-T7 platform with 100 bp paired-end reads.

### RNA-Seq raw data quality control, gene expression analysis, and immune cell **abundance estimation**

Raw sequencing data (raw reads) from MGISEQ-T7 was processed to filter out low-quality reads. Clean reads from each sample were aligned against the hg19 reference by STAR (v2.7.8) (26). The read counts and transcripts per million (TPM) values were calculated based on aligned RNA sequencing reads to reference transcripts downloaded from gencode (v27lift37) database, as implemented in rsem (v1.3.0) (27). Based on the gene expression matrix, R packages xCell (28) and web tool ImmunCellAI (29) were used to estimate the infiltration abundance of immune cells for each sample.

### T cell receptor sequencing and analysis

For T cell receptor (TCR) sequencing, total RNA was isolated from peripheral blood mononuclear cells (PBMCs) by RNeasy Plus Mini Kit (Qiagen, USA) according to the manufacturer’s instructions. Qubit™ RNA HS Assay Kit (ThermoFisher Scientific, USA) was applied to determine the final concentration. Then TCR β-chain of CDR3 was amplified by Multi-IR Library Prep Kit (iGeneTech, CHN). Sequencing procedures were utilized by the MGISEQ-T7 platform with 100-bp paired-end reads. Fastq reads were trimmed based on their low-quality 3’ ends to base. Trimmed pair-end reads were integrated according to overlapping alignment with the modified Needleman-Wunsch algorithm. MiXCR (v2.1.10) (30) was performed to identify the CDR3 sequences of V-D-J gene segments with reference sequences from the IMGT (31). VDJtools (v1.2.1) (32) was utilized to assess the immune repertoire sequencing. A frequency-based correction was performed on samples. The diversity of the TCR repertoire was calculated with the Shannon-Wiener index formula represents the frequency of clonotype i for the sample with n unique clonotypes. The evenness of the TCR repertoire was calculated with the formula , where H was Shannon Weiner index and S was the total number of species in a sample, across all samples in the dataset. Clonality is defined as a normalized Shannon index over n unique clones: clonality = 1 − H/ln(n). The D50 index was also performed to evaluate the diversity according to the top 50% of the total TCR clones in the samples. According to clonotype abundance, TCR clonotypes were defined as large clones when their frequency > 0.001. Based on the frequency ranking, the top 20 most frequent TCR clones and large clones are defined as the predominant TCR clones.

### Statistical analysis

All statistical analyses were implemented by R 4.0 software or SPSS 27.0. Medians and inter quartile range (IQRs) were provided for distributions of observation duration and time intervals. Wilcoxon’s Rank-Sum test was used to compare across independent groups. Wilcoxon’s Signed-Rank test was used for the pairwise comparison samples. All reported *P* values are two-sided, and *P* values less than 0.05 were considered statistically significant.

## Results

### Overview of the patient cohort

From May 2020 to November 2023, 50 patients diagnosed with NSCLC at stage IIA-IIIA were screened for eligibility, of which 38 received two cycles of modified neoadjuvant chemo-immunotherapy (**Fig. 1A**). Each cycle featured two phases, corresponding to chemotherapy and immunotherapy (**Fig. 1B**). Following modified neoadjuvant chemo-immunotherapy, 31 patients underwent R0 resection (**Figure 1A and Table S1**). The postoperative tissues were subjected to pathological evaluation. Samples from a representative patient (P2) who achieved pCR after neoadjuvant treatment exhibited morphological alterations, characterized by extracellular mucin pools and fibrosis (**Fig. 1C**). Computed tomography scans demonstrated the obliteration of lung lesions following neoadjuvant therapy (**Fig. 1D**). 74.2% (27/31) participants received the modified adjuvant chemo-immunotherapy (from time-point c3d0 to c4d21) (**Fig. 1A and B**; **Table S1**). Detailed treatment regimens are provided in **Table S2**.

**Fig. 1.**
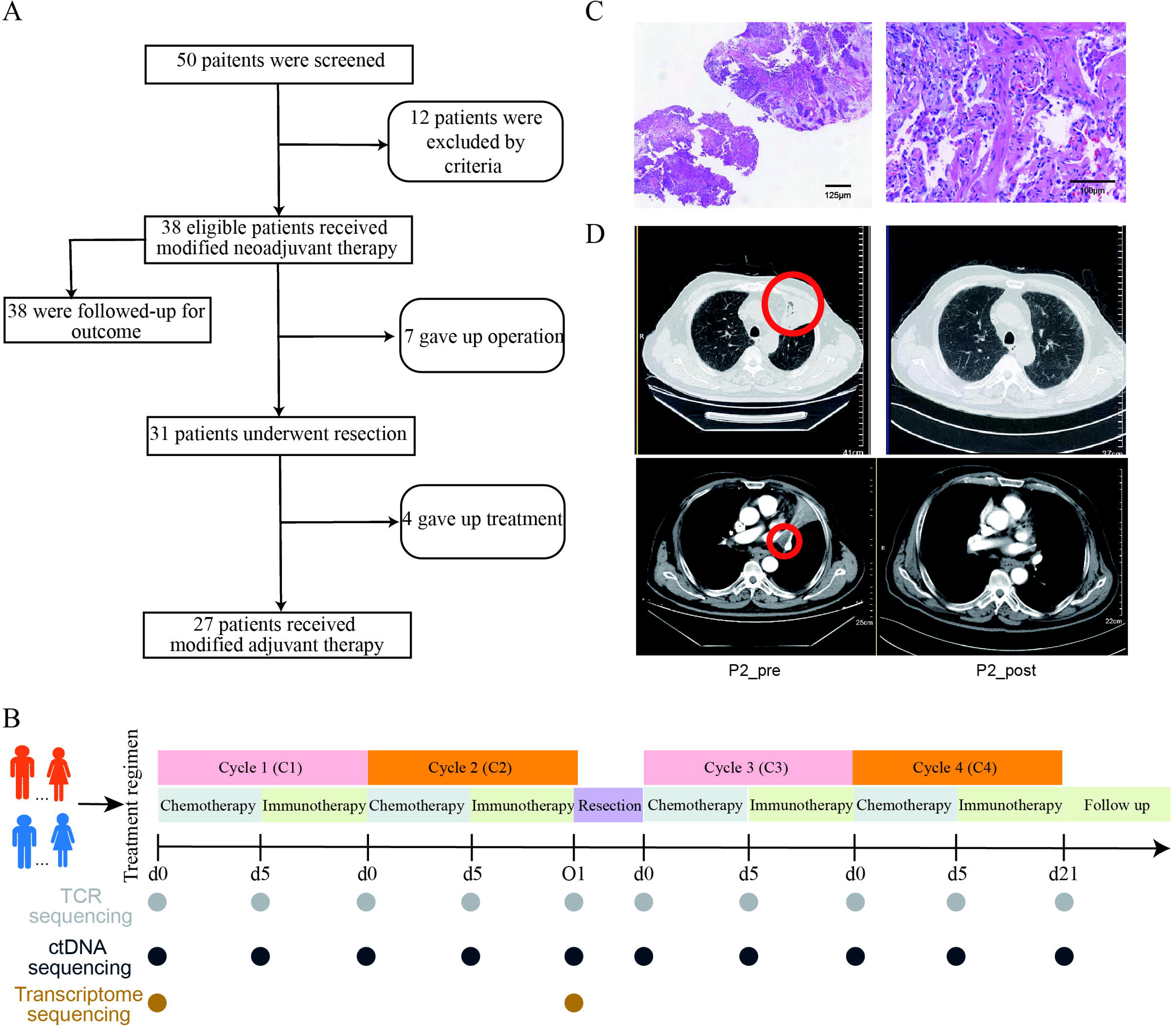
Overview of study design and representative images. **(A)** Flow diagram of the study. **(B)** Trial schema of the study. Patients with NSCLC received modified neoadjuvant chemo-immunotherapy, followed by surgical resection and modified adjuvant chemo-immunotherapy. Tumor tissues were collected at baseline and the surgery phase. Serial blood samples were collected at baseline and each time-point for TCR and ctDNA sequencing. **(C)** Representative hematoxylin and eosin staining of tumor specimens obtained from patient 2 (P2), who obtained pCR. **(D)** Representative computed tomographic imaging of lung lesions of P2. NSCLC, non-small cell lung cancer; TCR, T-cell receptor; ctDNA, circulating tumor DNA; pCR, pathologic complete response.

Most patients were men (78.7%) and had a smoking history (81.6%). The median age of patients was 64 years (range: 37-77). All patients had ECOG performance status of 0 or 1. Baseline patient characteristics are summarized in **Table 1** and detailed in **Table S1**.

**Table 1.**
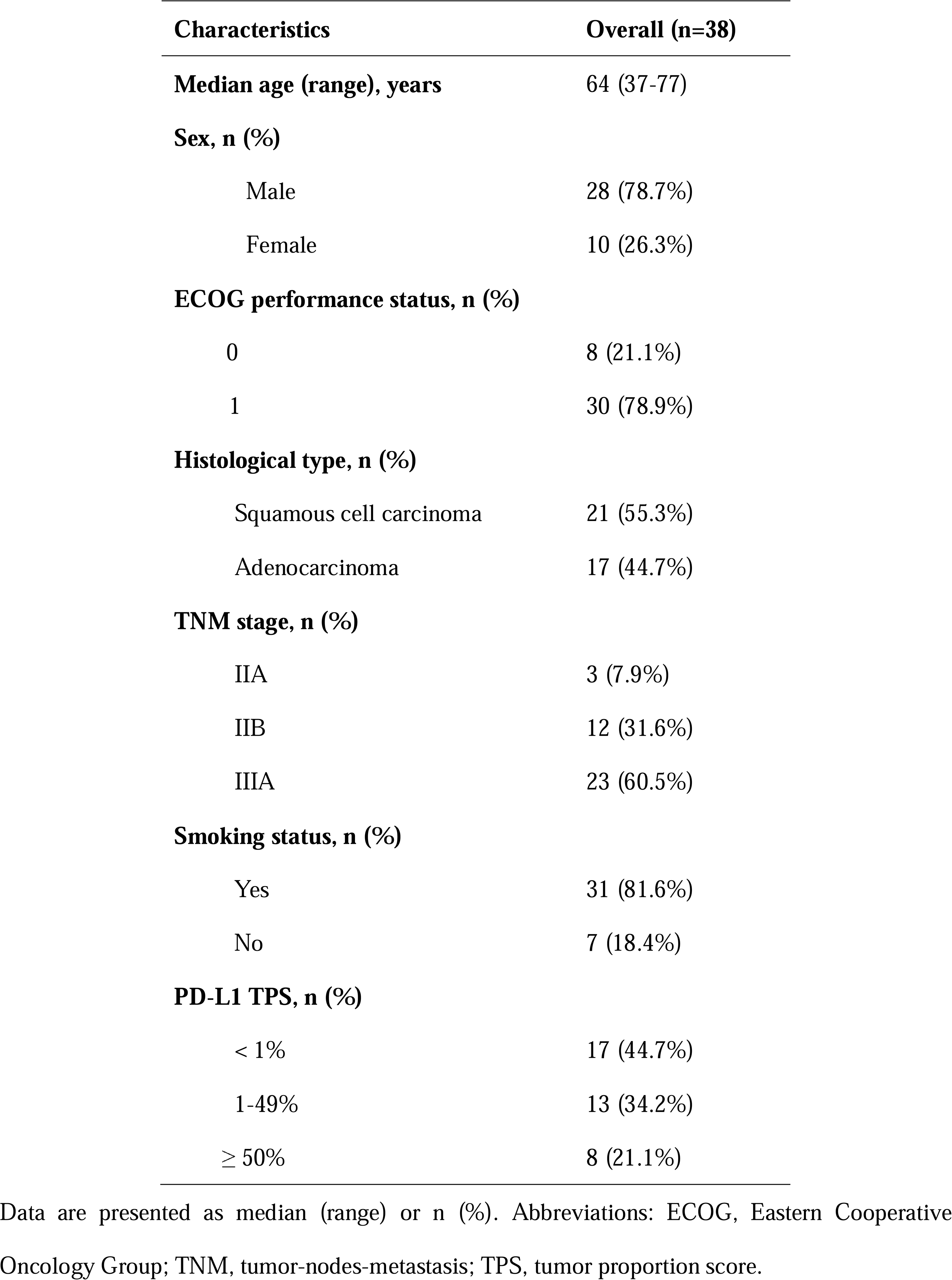
Baseline clinical characteristics of patients.

### Clinical efficacy and safety

After neoadjuvant therapy, 39.5% (15/38) patients achieved radiological downstaging (**Fig. 2A**; **Table 2**). The ratio of patients with CR, PR, and stable disease (SD) was 5.3% (2/38), 68.4% (26/38), and 26.3% (10/38), respectively, yielding an ORR of 73.7% (28/38) (**Fig. 2B**). 91.3% (21 of 23) were recurrence-free (**Fig. 2C**). The median interval between the last dose of sintilimab and surgery was 33 days (interquartile ranges (IQRs) 5.55 days) (**Table S1**). 58.1% (18/31) patients achieved a MPR, including 12 patients with a pCR, and 41.9% (13/31) patients achieved a non-MPR (**Fig. 2D**).

**Fig. 2.**
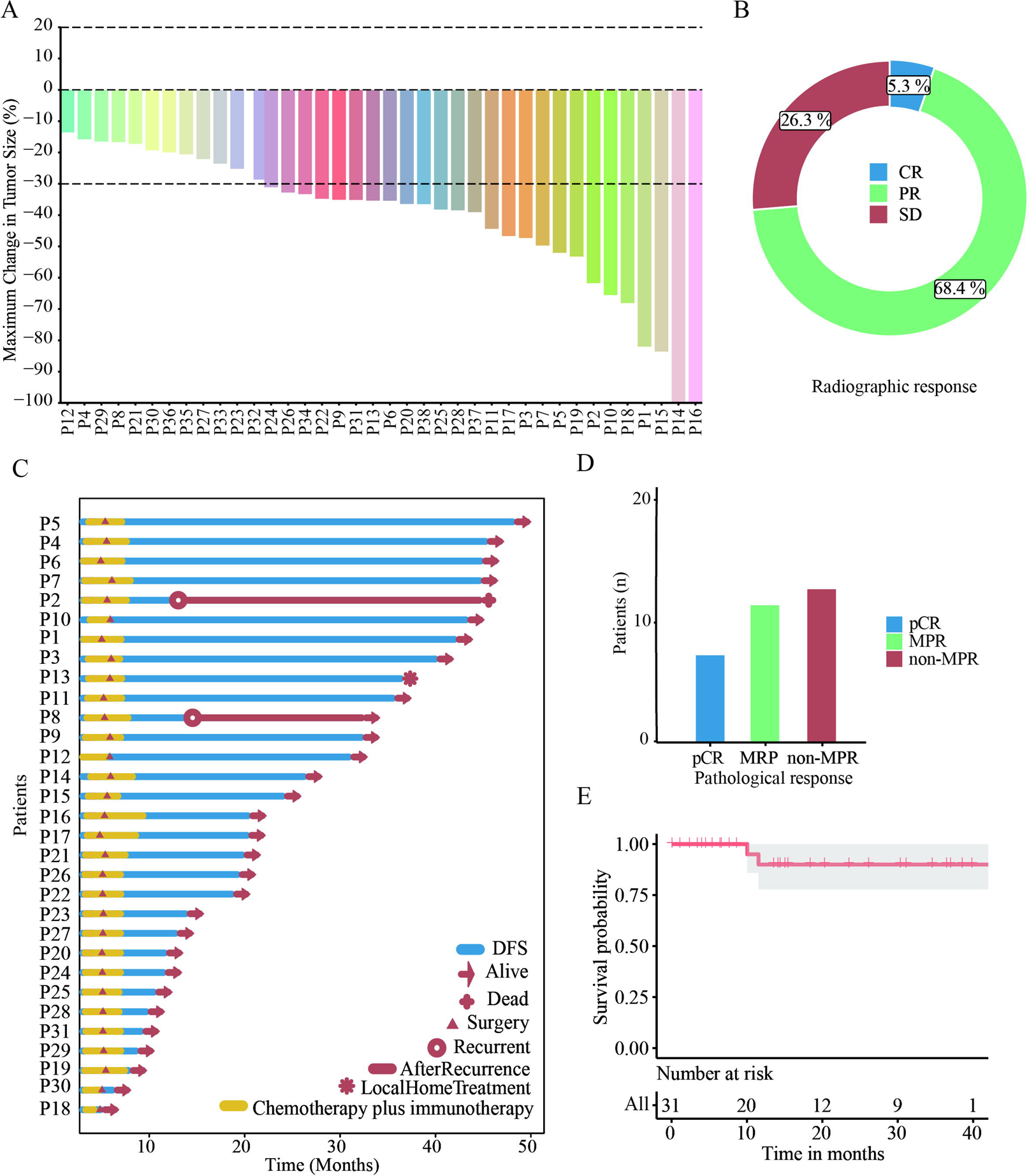
Clinical efficacy of modified neoadjuvant chemo-immunotherapy. **(A)** Tumor size changes from baseline based on radiological imaging. **(B)** Ring diagram showed the rate of overall response according to RECIST criteria (CR, PR, and SD) in patients. **(C)** Swimmer plot of 31 patients who underwent R0 resection involved in this trial. **(D)** Barplot showed the number of patients distributed in pathological response (pCR in blue, MPR in green, and non-MPR in red, respectively). **(E)** DFS curve for patients with modified treatment. CR, complete response; PR, partial response; SD, stable disease; pCR, completed pathological response; MPR, major pathological response; DFS, disease free survival.

**Table 2.** Response to modified neoadjuvant chemo-immunotherapy

| Response to Treatment | Rate (%), Patient No. |
| --- | --- |
| Tumor down-staging rate | 39.5% (15/38) |
| ORR | 73.7% (28/38) |
| R0 resection rate | 81.6% (31/38) |
| pCR rate | 38.7% (12/31) |
| MPR rate | 58.1% (18/31) |

Data are presented as n%. ORR, objective response rate; pCR, pathologic complete response;
MPR, major pathological response.

For patients receiving adjuvant therapy, the median interval between the surgery and the adjuvant therapy was 32.1 days (IQRs 7.2 days) (**Table S1**). Up to January 18, 2024, the median follow-up was 14.37 months (IQRs 24.22 months) (**Table S1**). The median OS are not reached (**Fig. 2E**). 44.7% (17/38) patients experienced at least one TRAE (**Table 3**). Grade 3 TRAEs occurred only in one patient. The most common grade 1 or 2 TRAEs were nausea (15.8%). Previously unreported toxic effects were not observed, and there were no instances of discontinuation of modified neoadjuvant chemo-immunotherapy due to toxicity.

**Table 3.** Summary of treatment-related adverse events

| **Treatment-related adverse events** | **Grade 1 or 2** | **Grade 3** |
| --- | --- | --- |
| **n (%)** | **16 (42.1%)** | **1 (2.6%)** |
| Nausea | 6 (15.8%) | 0 |
| Constipation | 3 (7.9%) | 0 |
| Decreased appetite | 3 (7.9%) | 0 |
| Anemia | 2 (5.3%) | 0 |
| Fever | 2 (5.3%) | 0 |
| Rash | 2 (5.3%) | 0 |
| Decreased neutrophil count | 2 (5.3%) | 0 |
| Diarrhea/colitis | 1 (2.6%) | 0 |
| Hepatotoxicity | 1 (2.6%) | 1 (2.6%) |
| Hypothyroidism | 1 (2.6%) | 0 |
| Adrenal insufficiency | 1 (2.6%) | 0 |
| Hyperthyroidism | 1 (2.6%) | 0 |
| Maculopapular | 0 | 0 |
| Hypersensitivity | 0 | 0 |
| Pneumonitis | 0 | 0 |
| Hypophysitis | 0 | 0 |
| thyroiditis | 0 | 0 |
| Diabetes mellitus | 0 | 0 |
| Neutropenia | 0 | 0 |

### Low-dose chemotherapy combined with delayed immunotherapy has a positive effect on T lymphocytes

To investigate whether dynamic changes in peripheral blood TCR repertoire were associated with treatment efficiency during modified chemo-immunotherapy, 49 peripheral blood samples were collected from serial time-points and evaluated using TCR sequencing (**Fig. 3A**). Since the predominant number of TCR clones is an important indicator of immune response and treatment efficacy (15, 33–35), the frequency of the top 20 TCR clones was analyzed at each treatment phase. Significant increase of top 20 TCR clones were observed in phase 1 to 3 (all *P* < 0.0001; **Fig. 3B**; **Fig. S1**). An overall increase of top 20 frequency was observed in the first cycle of neoadjuvant therapy (C1). In contrast, no significant changes were observed during adjuvant therapy (**Fig. 3C**; **Table S3**). These results suggested that the peripheral blood-derived top 20 TCR clones were phased increased not only during low-dose chemotherapy, but also during immunotherapy within neoadjuvant therapy.

**Fig. 3.**
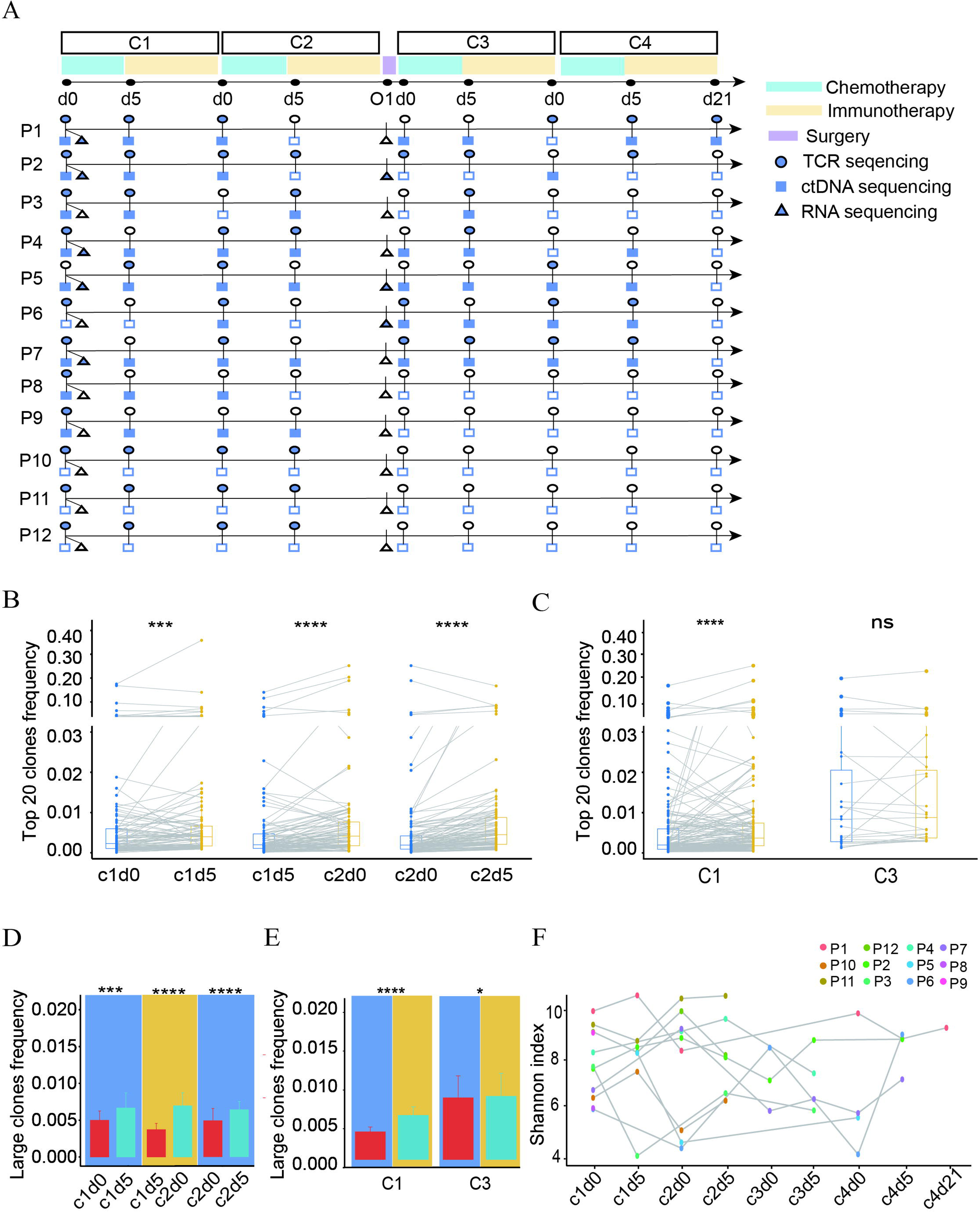
Dynamic changes in T cell repertoire during therapy. **(A)** The schematic of samples collection and analyses performed for each patient. Top boxes represent therapeutic phases, blue for chemotherapy, yellow for immunotherapy, and purple for operation. TCR sequencing is indicated as circle. Square and triangle represents ctDNA and RNA sequencing, respectively. Solid shapes are defined as available data, while open shapes are defined as unavailable data. **(B)** Comparison of top 20 TCR clones at different therapeutic phases. **(C)** Comparison of top 20 TCR clones at different therapeutic cycles. For boxplots, each box indicates the first quartile (Q1) and third quartile (Q3), and the black horizontal line represents the median; the upper whisker is the min[max(x), Q3 + 1.5 × IQR], and the lower whisker is the max[min(x), Q1 - 1.5 × IQR], where x represents the data, Q3 is the 75th percentile, Q1 is the 25th percentile, and IQR = Q3 - Q1. **(D)** Comparison of large TCR clones at different therapeutic phases. **(E)** Comparison of large TCR clones at different therapeutic cycles. Average and SD are shown. Wilcoxon’s Signed-Rank test was used for comparison in B-E (\**p* < 0.05, \*\**p* < 0.01, \*\*\**p* < 0.001, **** *p* < 0.0001). **(F)** Changes in diversity (Shannon index) of TCR clones for each patient. TCR, T-cell receptor; ctDNA, circulating tumor DNA; IQR, interquartile range.

According to clonotype abundance, the TCR clones were divided into 3 clonotypes, including large (frequency > 0.001), median (0.001-0.0001), and small (< 0.0001) clones. A markedly increase of large clones was observed during neoadjuvant therapy (all *P* < 0.05) (**Fig. 3D and E**), consistent with the trend of top 20 TCR clones. No significant difference was observed in the diversity, evenness, and clonality of TCR clones across the whole treatment procedure (**Fig. 3F**; **Fig. S2; Table S4**). The changes of T lymphocytes in the tumor microenvironment across the neoadjuvant therapy were analyzed based on transcriptomic data. As shown in **Figure S3**, the CD4+ and CD8+ T lymphocytes tended to increase during neoadjuvant therapy. Collectively, the neoadjuvant treatment had a positive effect on T lymphocytes.

### Reduced tumor burden in ctDNA was observed during neoadjuvant therapy

To monitor the dynamic changes of tumor burden in ctDNA during therapy, peripheral blood samples were collected at serial time-points (T0 to T7) and evaluated via ctDNA sequencing. The landscape of genomic alternations was shown in Figure 4A. Among the detected genes, *TP53* (50%) was the most frequently mutated gene, followed by *ARID1A* (11%), *DNMT3A* (11%), and *PIK3CA* (11%) (**Fig. 4A**; **Table S5**). Throughout the entire treatment, the maximal somatic variant allelic frequency (maxVAF) of ctDNA had a declining trend, especially in PH1 (T0 to T1) with dramatically decreased (*P* < 0.05) (**Fig. 4B**; **Fig. S4**), indicating that low-dose chemotherapy would induce tumor cell death thus reducing tumor burden. Notably, we observed that the changes in TCR and ctDNA exhibited opposite trends during the neoadjuvant therapy (**Fig.3B and 3D; Fig. 4B**). Taken together, these results suggest that low-dose chemotherapy combined with delayed immunotherapy not only cause the increase of tumor antigens shed into blood but also stimulate the body’s immunity to produce more tumor antigen-specific T lymphocytes.

**Fig. 4.**
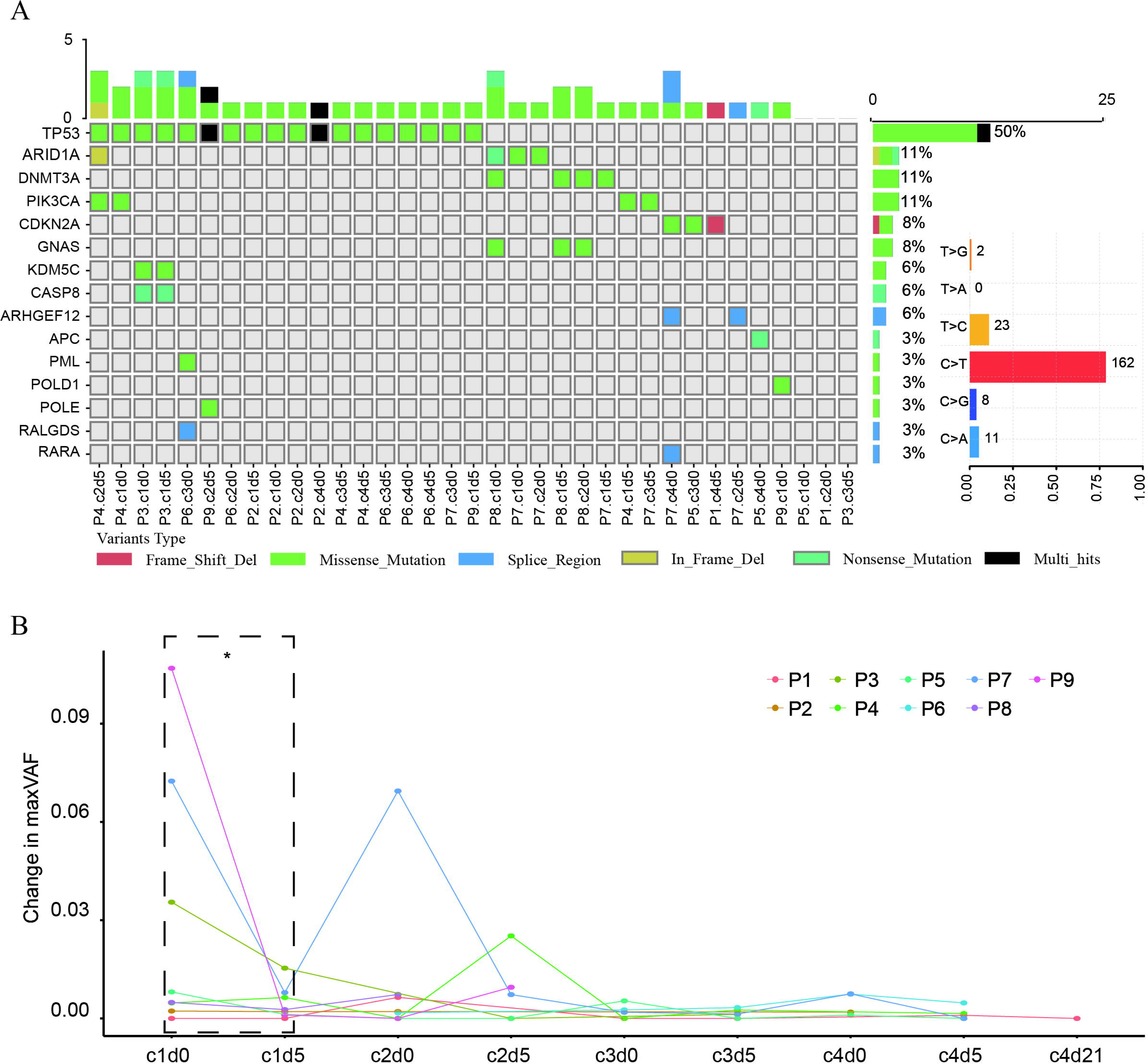
Characterization of ctDNA and the alteration of tumor burden during treatment. **(A)** Genomic landscape of mutations detected in plasma via ctDNA sequencing. Frequency of mutated genes were shown on the bar of middle and the proportion was shown in numerical form. Counts of mutations for each sample were displayed on top bar. Frequency for each of the six substitutions of the bases spectrum was shown on right. The scale lines below show the proportion of various mutation substitutions. **(B)** Changes in maxVAF across the entire course of therapy. Wilcoxon’s Signed-Rank test was used for comparison. (\**p* < 0.05). ctDNA, circulating tumor DNA; maxVAF, maximal somatic variant allelic frequency.

## Discussion

In recent clinical trials, the combination of neoadjuvant chemotherapy with immunotherapy by administering them concurrently in patients with resectable NSCLC has brought about longer event-free survival (6, 8, 36). Although chemotherapy promotes tumor cell death and T lymphocyte infiltration (17, 18), it may also have negative effects on T lymphocyte proliferation (37). Therefore, in the context of combination therapy, the dosage of chemotherapy and the timing of its administration relative to the other treatment need to be better adjusted to optimize the current therapeutic efficacy.

Our previous findings from both in vitro experiments and clinical trials have suggested that administering PD-1/PD-L1 inhibitors at 1-10 days (especially 3-5 days) after chemotherapy in lung cancer patients may be more effective than administering them before or concurrently (14). In this trial, we reported the safety and efficacy of low-dose platinum-based chemotherapy combined with delayed immunotherapy for patients with stage IIA to IIIA resectable NSCLC. After modified neoadjuvant therapy, ORR was achieved in 73.7% (28/38) of patients, which was higher than the report of CheckMate-816 (54%) (6) and comparable to those of NADIM (76%) (8) and LungMate 002 (76%) (38) trials. MPR was achieved in 47.4% (18/38) patients, with 31.6% (12/38) achieving pCR. During a median follow-up of 14.37 months, 91.3% (21/23) patients were recurrence-free. These results indicated that modified neoadjuvant chemo-immunotherapy was effective in patients with resectable NSCLC. Only 2.6% (1/38) patients experienced serious TRAEs after receiving modified neoadjuvant treatment, which is lower than the rates reported in CheckMate 816 (33.5%), NAMID (30.0%) (8), and LungMate 002 (6.0%) (38) trials. The reduction of chemotherapeutics may contribute to the low incidence of serious TRAEs. Additionally, we did not observe treatment discontinuation due to toxicity. These results provide evidences that low-dose platinum-based chemotherapy combined with delayed immunotherapy as a neoadjuvant therapy is safe and less toxic.

Previous research has illustrated an association between clinical benefit and TCR repertoire as well as ctDNA level at baseline (39, 40). However, the dynamic changes in the TCR repertoire and ctDNA level during neoadjuvant treatment have been less studied. In our study, we found that the predominant TCR clones were greatly increased after the treatment of chemotherapy from phase 1 to 3, indicating that low-dose platinum-based chemotherapy did not inhibit the proliferation of predominant T cell clones which create a beneficial microenvironment for subsequent immunotherapy. Moreover, ICI treatment significantly increased the number of predominant TCR clones during neoadjuvant therapy, suggesting that immunotherapy has a positive impact on TCR clones, consistent with the study by Forde et al (41). During adjuvant therapy, no significant fluctuation in the predominant TCR clones was observed, probably because the tumor lesions were cleared. Under such circumstances, no tumor antigen was released, thus no tumor antigen-specific T cells were proliferated. Similarly, no significant change was observed in evenness or clonality of TCR. MaxVAF of ctDNA was dramatically dropped during phase 1, which is accordance with previous studies (42–44). Notably, the maxVAF of ctDNA and predominant TCR clones showed opposite trends of change in phase 1. Moreover, the maxVAF of ctDNA was close to 0 during adjuvant therapy stage.

Our study had several limitations that need to be acknowledged. First, it was a pilot study with a relatively small cohort and lacked a control arm. Therefore, validation of our findings in larger cohorts is necessary. Second, the follow-up period was short, and longer-term results are needed to investigate whether modified therapy provides long-term survival benefits. Finally, innovative techniques should be incorporated to further explore the underlying mechanisms of superior clinical efficacy and reduced TRAEs.

In this study, the modified neoadjuvant low-dose platinum-based chemotherapy combined with delayed immunotherapy in patients with resectable NSCLC exhibited promising efficacy and feasibility. The multi-omics analyses provide the dynamic trajectories of the TCR repertoire and tumor burden, which can aim in better understanding the mechanisms of superior efficiency and the feasibility of modified chemo-immunotherapy. In the future, it is possible to conduct large-scale clinical research using this modified chemo-immunotherapy.

## Acknowledgements

We would like to thank Innovent Biologics, Inc for providing sintilimab for this study.

## Author’s contributions

C.Y. Liang: Conceptualization, Formal analysis, Methodology, Writing - original draft. Q. Song: Formal analysis, Methodology, Funding acquisition, Writing - original draft. W.H. Zhou and N. Li: Formal analysis, Methodology, Writing - original draft. C.H. Pan, Q. Xiong B. Yang, and X.L Zhang: Formal analysis. S.H Zhao, W.W. Shi, X. Yan, J.T. Guo, S.J. Sun, Z.H. Dong, and Y.P. Long: Validation, Writing - reviewing & editing. B. Yang, J. Li, Y. Hu, and H.T. Luo: Conceptualization, Investigation, Validation, Verification of underlying data, Supervision, Writing - reviewing & editing.

## Data availability statement

All the analysis data can be found in the supplementary tables. The dataset supporting the conclusions of this article is available in the Genome Sequence Archive for Human (GSA-H) repository with accession number (HRA002904) and can be accessed thru the URL (BIG Search - National Genomics Data Center - BIG Search (cncb.ac.cn)) after reasonable request.

## Declarations

### Ethics approval and consent to participate

This study was conducted according to the guidelines of the Ethics Committee of the General Hospital of the People’s Liberation Army (S2020-144-01) and written informed consent was obtained from all participants.

## Consent for publication

Not applicable.

## Supplementary materials

**Fig. S1 Dynamic change of top 20 TCR clones in the distinct phases.**

**(A)** Changes of top 20 TCR clones of P1, P2, P3, P10, P11, and P12 in T0 to T1 (PH 1). **(B)** Changes of top 20 TCR clones of P1, P2, P5, P10, P11, and P12 in T1 to T2 (PH 2). **(C)** Changes of top 20 TCR clones of P2, P4, P10, P11, and P12 in T2 to T3 (PH 3). For boxplots, each box indicates the first quartile (Q1) and third quartile (Q3), and the black horizontal line represents the median; the upper whisker is the min[max(x), Q3 + 1.5 × IQR], and the lower whisker is the max[min(x), Q1 - 1.5 × IQR], where x represents the data, Q3 is the 75th percentile, Q1 is the 25th percentile, and IQR = Q3 - Q1. Wilcoxon’s Signed-Rank test was used for comparison (\**p* < 0.05, \*\**p* < 0.01, \*\*\**p* < 0.001, **** *p* < 0.0001). TCR, T-cell receptor; IQR, interquartile range.

**Fig. S2 Characterization of temporal dynamics of the T cell repertoire in the longitudinal blood.**

**(A)** Changes of the D50 index among patients. **(B)** Changes of Pielou index (evenness) among patients. **(C)** Changes of clonality index among patients.

**Fig. S3 Infiltration of CD4+ and CD8+ T lymphocytes after modified neoadjuvant therapy.**

**(A)** Relative proportion of lymphocytes was assessed by ImmunCellAI. **(B)** Relative proportion of lymphocytes was assessed by xCell. For boxplots, each box indicates the first quartile (Q1) and third quartile (Q3), and the black horizontal line represents the median; the upper whisker is the min[max(x), Q3 + 1.5 × IQR], and the lower whisker is the max[min(x), Q1 - 1.5 × IQR], where x represents the data, Q3 is the 75th percentile, Q1 is the 25th percentile, and IQR = Q3 - Q1. Wilcoxon’s Signed-Rank test was used for comparison. Tcm, central memory T cells; Tem, Effector memory T cells; IQR, interquartile range.

**Fig. S4 Change in maxVAF across different phases.** For boxplots, each box indicates the first quartile (Q1) and third quartile (Q3), and the black horizontal line represents the median; the upper whisker is the min[max(x), Q3 + 1.5 × IQR], and the lower whisker is the max[min(x), Q1 - 1.5 × IQR], where x represents the data, Q3 is the 75th percentile, Q1 is the 25th percentile, and IQR = Q3 - Q1. Wilcoxon’s Signed-Rank test was used for comparison. IQR, interquartile range.

**Table S1. Clinical characteristics of patients**

**Table S2. List of treatment regimens**

**Table S3. List of top 20 TCR clones frequency across the whole modified therapy**

**Table S4. TCR characteristics across the whole modified therapy**

**Table S5. T cells abundance estimated by xCell**

**Table S6. T cells abundance estimated by ImmunCellAI**

**Table S7. Annotation of ctDNA somatic mutations**

