## Supplemental figures for "Low-dose chemotherapy combined with delayed immunotherapy in the neoadjuvant treatment of non-small cell lung cancer and dynamic monitoring of the drug response in peripheral blood"

Supplementary Figures

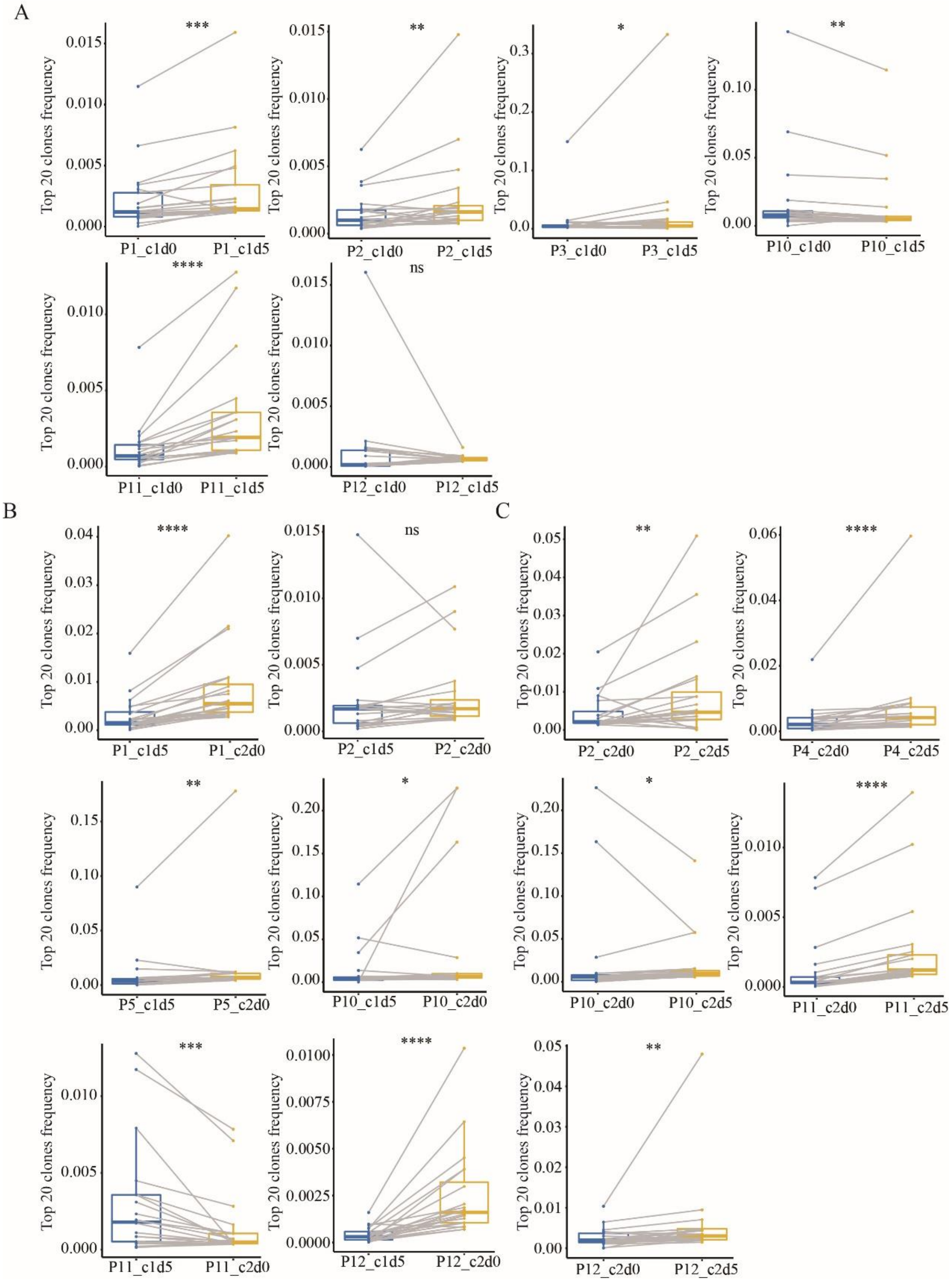


**Supplementary Figure. S1 Dynamic change oftop 20 TCR clones dynamic changes in the different stages per sample. A.** Changes of top20 TCR clones of P1, P2, P3, P10, P11, and P12 in c1d0 to c1d5. **B.** Changes of top20 TCR clones of P1, P2, P5, P10, P11, and P12 in c1d5 to c2d0. **C.** Changes of top20 TCR clones of P2, P4, P10, P11, and P12 in c2d0 to c2d5. For boxplots, each box indicates the first quartile (Q1) and third quartile (Q3), and the black horizontal line represents the median; the upper whisker is the min[max(x), Q3 + 1.5 × IQR], and the lower whisker is the max[min(x), Q1 -1.5 × IQR], where x represents the data, Q3 is the 75th percentile, Q1 is the 25th percentile, and IQR = Q3 -Q1. Wilcoxon's Signed-Rank test was used for comparison (*$p < 0.05$, **$p < 0.01$, ***$p < 0.001$, **** $p < 0.0001$).TCR, T-cell receptor; IQR, interquartile range.

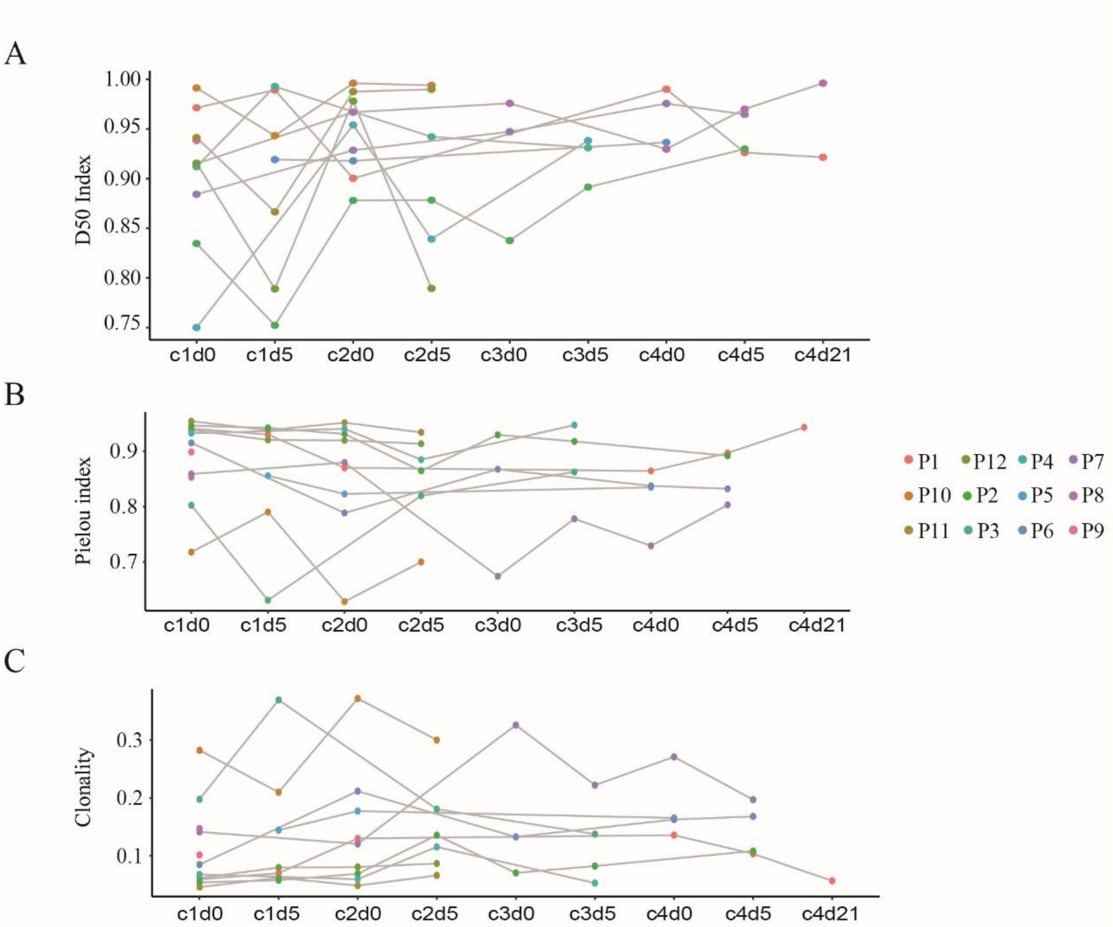


**Supplementary Figure. S2 Characterization of temporal dynamics of the T cell repertoire in the longitudinal blood among patients. A.** Changes of the D50 index among patients. **B.** Changes of Pielou index (evenness) among patients. **C.** Changes of clonality index among patients.

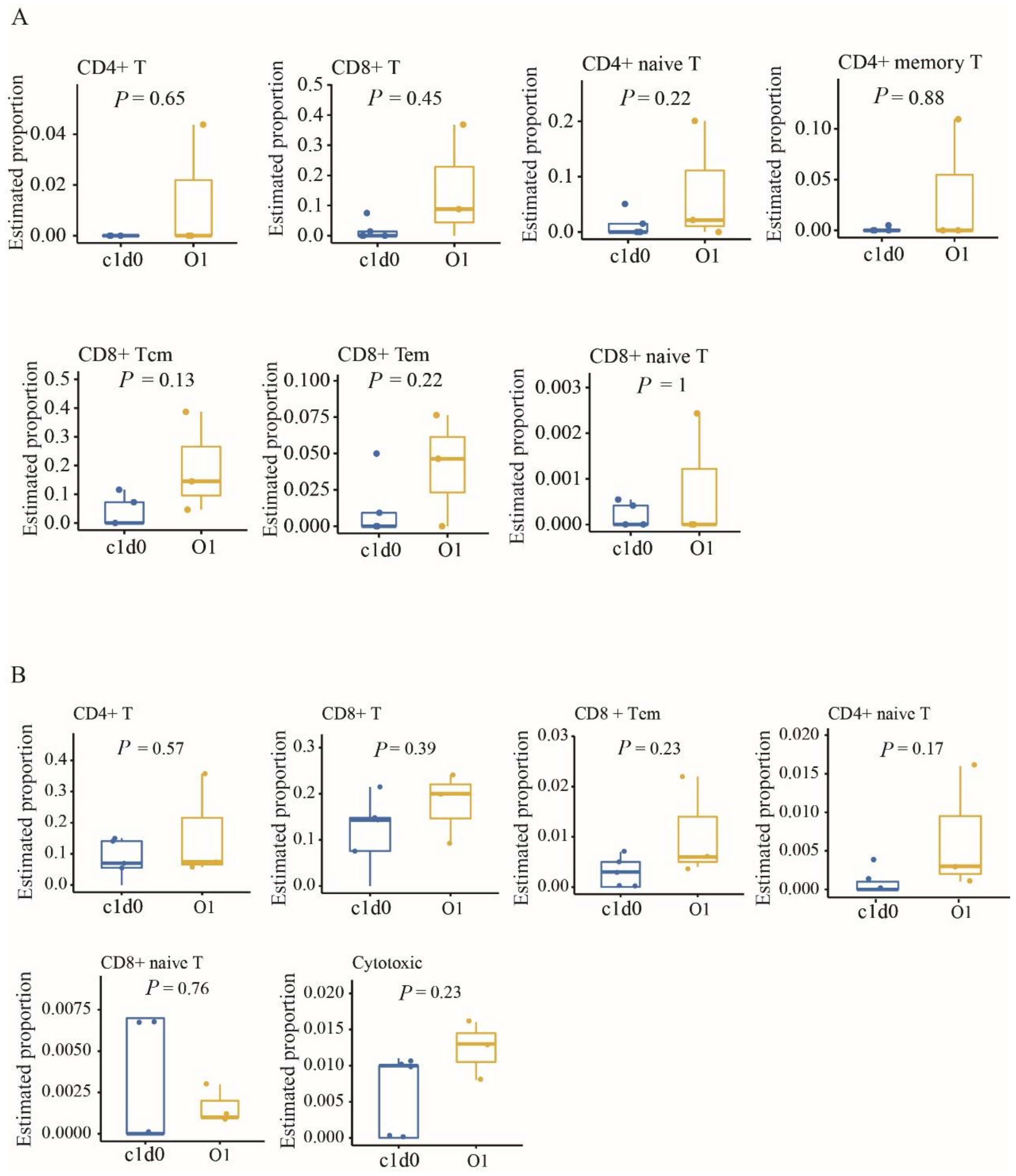


**Supplementary Figure. S3 Infiltration of CD4+ and CD8+ T lymphocytes after modified neoadjuvant therapy. A.** Relative proportion of lymphocytes was assessed by ImmunCellAI. **B.** Relative proportion of lymphocytes was assessed by xCell. For boxplots, each box indicates the first quartile (Q1) and third quartile (Q3), and the black horizontal line represents the median; the upper whisker is the min[max(x), Q3 + 1.5 × IQR], and the lower whisker is the max[min(x), Q1 -1.5 × IQR], where x represents the data, Q3 is the 75th percentile, Q1 is the 25th percentile, and IQR = Q3 -Q1. Wilcoxon's Signed-Rank test was used for comparison. Tcm, central memory T cells; Tem, Effector memory T cells; IQR, interquartile range.

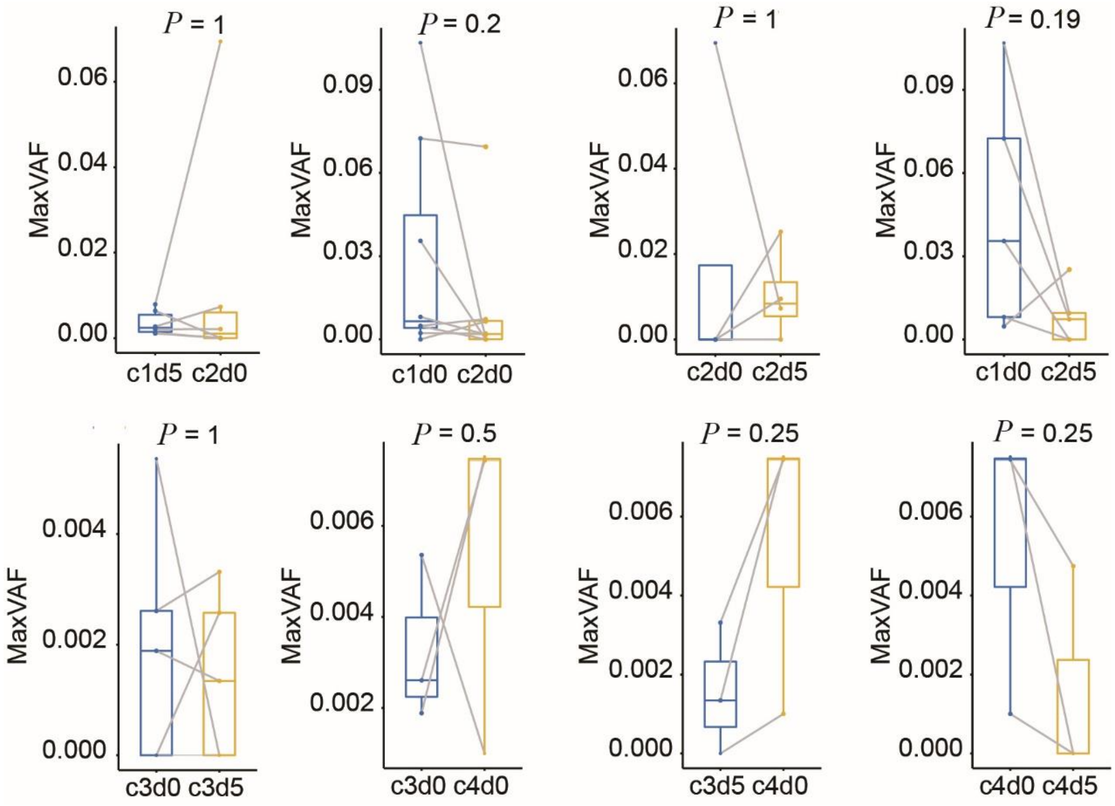


**Supplementary Figure. S4 Change in maxVAF across different phases.** For boxplots, each box indicates the first quartile (Q1) and third quartile (Q3), and the black horizontal line represents the median; the upper whisker is the min[max(x), Q3 + 1.5 × IQR], and the lower whisker is the max[min(x), Q1 -1.5 × IQR], where x represents the data, Q3 is the 75th percentile, Q1 is the 25th percentile, and IQR = Q3 -Q1. Wilcoxon's Signed-Rank test was used for comparison. IQR, interquartile range.
