## Supplementary Table 1 for "Low-dose chemotherapy combined with delayed immunotherapy in the neoadjuvant treatment of non-small cell lung cancer and dynamic monitoring of the drug response in peripheral blood"

### Table S1. Clinical characteristics of patients

| Patient | Gender | Age ranges | Histological type | radiographic evaluation | Histological group |
|---|---|---|---|---|---|
| P1 | F | 51-55 | adenocarcinoma | CR | PCR |
| P2 | M | 71-75 | adenocarcinoma | PR | PCR |
| P3 | M | 51-55 | squamous | PR | PCR |
| P4 | M | 66-70 | squamous | PR | MPR |
| P5 | M | 56-60 | adenocarcinoma | PR | non-MPR |
| P6 | M | 66-70 | squamous | PR | MPR |
| P7 | M | 66-70 | squamous | PR | non-MPR |
| P8 | F | 61-65 | adenocarcinoma | PR | non-MPR |
| P9 | M | 61-65 | squamous | PR | non-MPR |
| P10 | M | 76-80 | squamous | PR | MPR |
| P11 | F | 66-70 | squamous | PR | MPR |
| P12 | M | 71-75 | adenocarcinoma | PR | non-MPR |
| P13 | M | 56-60 | adenocarcinoma | PR | MPR |
| P14 | M | 71-75 | squamous | CR | PCR |
| P15 | M | 61-65 | squamous | PR | PCR |
| P16 | M | 56-60 | squamous | PR | PCR |
| P17 | M | 46-50 | squamous | PR | MPR |
| P18 | M | 66-70 | squamous | PR | PCR |
| P19 | M | 36-40 | squamous | PR | PCR |
| P20 | M | 51-55 | squamous | SD | non-MPR |
| P21 | M | 66-70 | squamous | PR | PCR |
| P22 | M | 56-60 | squamous | SD | non-MPR |
| P23 | F | 66-70 | adenocarcinoma | SD | non-MPR |
| P24 | M | 61-65 | adenocarcinoma | SD | non-MPR |
| P25 | M | 61-65 | adenocarcinoma | SD | non-MPR |
| P26 | F | 76-80 | adenocarcinoma | PR | PCR |
| P27 | M | 66-70 | adenocarcinoma | SD | non-MPR |
| P28 | M | 71-75 | adenocarcinoma | SD | non-MPR |
| P29 | F | 56-60 | adenocarcinoma | PR | PCR |
| P30 | F | 71-75 | adenocarcinoma | PR | PCR |
| P31 | M | 56-60 | adenocarcinoma | SD | non-MPR |
| P32 | M | 71-75 | squamous | PR | unkown |
| P33 | F | 71-75 | squamous | PR | unkown |
| P34 | M | 71-75 | squamous | SD | unkown |
| P35 | M | 61-65 | squamous | SD | unkown |
| P36 | F | 61-65 | squamous | SD | unkown |
| P37 | F | 71-75 | adenocarcinoma | PR | unkown |
| P38 | M | 61-65 | adenocarcinoma | SD | unkown |

| Pathological diagnosis time | Neoadjuvant therapy start time （C1） | Neoadjuvant therapy start time （C2） | Surgery time | Adjuvant therapy start time(C3) |
|---|---|---|---|---|
| 11-2020 | 11-2020 | 12-2020 | 1-2021 | 2-2021 |
| 8-2020 | 8-2020 | 9-2020 | 11-2020 | 12-2020 |
| 1-2021 | 1-2021 | 2-2021 | 3-2021 | 5-2021 |
| 7-2020 | 8-2020 | 8-2020 | 11-2020 | 11-2020 |
| 5-2020 | 5-2020 | 6-2020 | 7-2020 | 8-2020 |
| 8-2020 | 8-2020 | 9-2020 | 10-2020 | 11-2020 |
| 8-2020 | 8-2020 | 9-2020 | 11-2020 | 12-2020 |
| 8-2021 | 9-2021 | 9-2021 | 11-2021 | 12-2021 |
| 8-2021 | 9-2021 | 9-2021 | 11-2021 | 12-2021 |
| 9-2020 | 10-2020 | 11-2020 | 1-2021 | |
| 5-2021 | 6-2021 | 6-2021 | 7-2021 | 8-2021 |
| 10-2021 | 12-2021 | 1-2022 | 2-2022 | |
| 4-2021 | 5-2021 | 6-2021 | 6-2021 | 8-2021 |
| 2-2022 | 3-2022 | 4-2022 | 5-2022 | 6-2022 |
| 4-2022 | 5-2022 | 6-2022 | 7-2022 | 8-2022 |
| 8-2022 | 8-2022 | 9-2022 | 10-2022 | 1-2023 |
| 8-2022 | 9-2022 | 9-2022 | 10-2022 | 12-2022 |
| 11-2023 | 12-2023 | 12-2023 | 1-2024 | |
| 8-2023 | 9-2023 | 9-2023 | 11-2023 | 12-2023 |
| 5-2023 | 5-2023 | 6-2023 | 7-2023 | 8-2023 |
| 9-2022 | 9-2022 | 10-2022 | 11-2022 | 12-2022 |
| 10-2022 | 10-2022 | 11-2022 | 12-2022 | 1-2023 |
| 3-2023 | 3-2023 | 4-2023 | 5-2023 | 6-2023 |
| 5-2023 | 5-2023 | 6-2023 | 7-2023 | 8-2023 |
| 6-2023 | 6-2023 | 7-2023 | 8-2023 | 9-2023 |
| 9-2022 | 9-2022 | 10-2022 | 11-2022 | 12-2022 |
| 4-2023 | 4-2023 | 5-2023 | 6-2023 | 7-2023 |
| 7-2023 | 7-2023 | 8-2023 | 9-2023 | 10-2023 |
| 8-2023 | 8-2023 | 9-2023 | 10-2023 | 11-2023 |
| 10-2023 | 10-2023 | 11-2023 | 12-2023 | |
| 7-2023 | 7-2023 | 8-2023 | 9-2023 | 10-2023 |
| 9-2020 | 9-2020 | 10-2020 | | |
| 11-2020 | 11-2020 | 12-2020 | | |
| 6-2021 | 6-2021 | 7-2021 | | |
| 6-2021 | 8-2021 | 8-2021 | | |
| 7-2021 | 7-2021 | 8-2021 | | |
| 8-2021 | 9-2021 | 9-2021 | | |
| 6-2023 | 6-2023 | 7-2023 | | |

| Adjuvant therapy start time（C4） | OS status | PFS status | Progression or last follow up |
|---|---|---|---|
| 3-2021 | F | F | 1-2025 |
| 1-2021 | T | T | 11-2021 |
| | F | T | 4-2022 |
| 12-2020 | F | F | 1-2025 |
| 8-2020 | F | F | 1-2025 |
| 12-2020 | F | F | 1-2025 |
| 1-2021 | F | F | 1-2025 |
| 1-2022 | F | F | 1-2025 |
| | F | F | 1-2025 |
| | F | F | 1-2025 |
| 9-2021 | F | F | 1-2025 |
| | F | F | 1-2025 |
| | F | F | 1-2025 |
| 7-2022 | F | F | 1-2025 |
| | F | F | 1-2025 |
| 2-2023 | F | F | 1-2025 |
| 2-2023 | F | T | 5-2023 |
| | F | F | 1-2025 |
| 1-2024 | F | F | 1-2025 |
| 9-2023 | F | F | 1-2025 |
| 1-2023 | F | F | 1-2025 |
| 2-2023 | F | F | 1-2025 |
| 6-2023 | F | F | 1-2025 |
| 9-2023 | F | F | 1-2025 |
| 10-2023 | F | F | 1-2025 |
| 1-2023 | F | F | 1-2025 |
| 7-2023 | F | F | 1-2025 |
| 10-2023 | F | F | 1-2025 |
| 12-2023 | F | F | 1-2025 |
| | F | F | 1-2025 |
| 11-2023 | F | F | 1-2025 |
