## Supplemental Table 2 for "Low-dose chemotherapy combined with delayed immunotherapy in the neoadjuvant treatment of non-small cell lung cancer and dynamic monitoring of the drug response in peripheral blood"

**Table S2. List of treatment regimens**

| Patients | Neoadjuvant treatment | Neoadjuvant treatment cycles |
|---|---|---|
| P1 | pemetrexed + cis-platinum+Sintilimab | 2 |
| P2 | pemetrexed + cis-platinum+Sintilimab | 2 |
| P3 | Albumin-bound paclitaxel + cis-platinum+Sintilimab | 2 |
| P4 | Albumin-bound paclitaxel + cis-platinum+Sintilimab | 2 |
| P5 | pemetrexed + cis-platinum+Sintilimab | 2 |
| P6 | Albumin-bound paclitaxel + cis-platinum+Sintilimab | 2 |
| P7 | Albumin-bound paclitaxel + cis-platinum+Sintilimab | 2 |
| P8 | pemetrexed + cis-platinum+Sintilimab | 2 |
| P9 | Albumin-bound paclitaxel + cis-platinum+Sintilimab | 2 |
| P10 | Albumin-bound paclitaxel + cis-platinum+Sintilimab | 2 |
| P11 | Albumin-bound paclitaxel + cis-platinum+Sintilimab | 2 |
| P12 | pemetrexed + cis-platinum+Sintilimab | 2 |
| P13 | pemetrexed + cis-platinum+Sintilimab | 2 |
| P14 | Albumin-bound paclitaxel + cis-platinum+Sintilimab | 2 |
| P15 | Albumin-bound paclitaxel + cis-platinum+Sintilimab | 2 |
| P16 | Albumin-bound paclitaxel + cis-platinum+Sintilimab | 2 |
| P17 | Albumin-bound paclitaxel + cis-platinum+Sintilimab | 2 |
| P18 | Albumin-bound paclitaxel + cis-platinum+Sintilimab | 2 |
| P19 | Albumin-bound paclitaxel + cis-platinum+Sintilimab | 2 |
| P20 | Albumin-bound paclitaxel + cis-platinum+Sintilimab | 2 |
| P21 | Albumin-bound paclitaxel + cis-platinum+Sintilimab | 2 |
| P22 | Albumin-bound paclitaxel + cis-platinum+Sintilimab | 2 |
| P23 | pemetrexed + cis-platinum+Sintilimab | 2 |
| P24 | pemetrexed + cis-platinum+Sintilimab | 2 |
| P25 | pemetrexed + cis-platinum+Sintilimab | 2 |
| P26 | pemetrexed + cis-platinum+Sintilimab | 2 |
| P27 | pemetrexed + cis-platinum+Sintilimab | 2 |
| P28 | pemetrexed + cis-platinum+Sintilimab | 2 |
| P29 | pemetrexed + cis-platinum+Sintilimab | 2 |
| P30 | pemetrexed + cis-platinum+Sintilimab | 2 |
| P31 | pemetrexed + cis-platinum+Sintilimab | 2 |
| P32 | Albumin-bound paclitaxel + cis-platinum+Sintilimab | 2 |
| P33 | Albumin-bound paclitaxel + cis-platinum+Sintilimab | 2 |
| P34 | Albumin-bound paclitaxel + cis-platinum+Sintilimab | 2 |
| P35 | Albumin-bound paclitaxel + cis-platinum+Sintilimab | 2 |
| P36 | Albumin-bound paclitaxel + cis-platinum+Sintilimab | 2 |
| P37 | pemetrexed + cis-platinum+Sintilimab | 2 |
| P38 | pemetrexed + cis-platinum+Sintilimab | 2 |

| Adjuvant treatment | Adjuvant treatment cycles |
|---|---|
| pemetrexed + cis-platinum+Sintilimab | 2 |
| pemetrexed + cis-platinum+Sintilimab | 2 |
| Albumin-bound paclitaxel + cis-platinum+Sintilimab | 1 |
| Albumin-bound paclitaxel + cis-platinum+Sintilimab | 2 |
| pemetrexed + cis-platinum+Sintilimab | 2 |
| Albumin-bound paclitaxel + cis-platinum+Sintilimab | 2 |
| Albumin-bound paclitaxel + cis-platinum+Sintilimab | 2 |
| pemetrexed + cis-platinum+Sintilimab | 2 |
| Albumin-bound paclitaxel + cis-platinum+Sintilimab | 1 |
| / | 0 |
| Albumin-bound paclitaxel + cis-platinum+Sintilimab | 2 |
| / | 0 |
| pemetrexed + cis-platinum+Sintilimab | 1 |
| Albumin-bound paclitaxel + cis-platinum+Sintilimab | 2 |
| Albumin-bound paclitaxel + cis-platinum+Sintilimab | 1 |
| Albumin-bound paclitaxel + cis-platinum+Sintilimab | 2 |
| Albumin-bound paclitaxel + cis-platinum+Sintilimab | 2 |
| / | 0 |
| Albumin-bound paclitaxel + cis-platinum+Sintilimab | 2 |
| Albumin-bound paclitaxel + cis-platinum+Sintilimab | 2 |
| Albumin-bound paclitaxel + cis-platinum+Sintilimab | 2 |
| Albumin-bound paclitaxel + cis-platinum+Sintilimab | 2 |
| pemetrexed + cis-platinum+Sintilimab | 2 |
| pemetrexed + cis-platinum+Sintilimab | 2 |
| pemetrexed + cis-platinum+Sintilimab | 2 |
| pemetrexed + cis-platinum+Sintilimab | 2 |
| pemetrexed + cis-platinum+Sintilimab | 2 |
| pemetrexed + cis-platinum+Sintilimab | 2 |
| pemetrexed + cis-platinum+Sintilimab | 2 |
| / | 0 |
| pemetrexed + cis-platinum+Sintilimab | 2 |
| / | 0 |
| / | 0 |
| / | 0 |
| / | 0 |
| / | 0 |
| / | 0 |
| / | 0 |
