## Supplemental Table 3 for "Low-dose chemotherapy combined with delayed immunotherapy in the neoadjuvant treatment of non-small cell lung cancer and dynamic monitoring of the drug response in peripheral blood"

| Table S3. List of top 20 TCR clones frequency across the whole modified therapy | | | |
|---|---|---|---|
| **Samples** | **Cdr3aa** | **Frequency** | **Time point** |
| P10T0 | CAISDRIYGYTF | 0.006172227 | T0 |
| P3T0 | CASDSGLADTQYF | 0.004044137 | T0 |
| P11T0 | CASIPEEGRTQYF | 0.000613675 | T0 |
| P1T0 | CASKLGAAGLFF | 0.002769107 | T0 |
| P2T0 | CASMVRYTGELFF | 0.003863406 | T0 |
| P2T0 | CASRGPTKGSSYEQYF | 0.000429532 | T0 |
| P1T0 | CASRGTSGIYEQYF | 0.01148744 | T0 |
| P10T0 | CASRIPGRRTDTQYF | 0.010436401 | T0 |
| P2T0 | CASRLHPGDTQYF | 0.006254163 | T0 |
| P2T0 | CASRPATGGDFYGYTF | 0.000727855 | T0 |
| P10T0 | CASRPRDRYQETQYF | 0.01877756 | T0 |
| P10T0 | CASSAVTGLAGGPYEQYF | 0.005963929 | T0 |
| P11T0 | CASSDGTGFYGYTF | 0.001383034 | T0 |
| P1T0 | CASSDSTSGGTLPQYF | 0.003421364 | T0 |
| P3T0 | CASSEDYGREETQYF | 0.003867851 | T0 |
| P10T0 | CASSEEPGGIEQYF | 0.005832731 | T0 |
| P10T0 | CASSELAGGPETQYF | 0.009306434 | T0 |
| P2T0 | CASSFAAVLSMNTEAFF | 0.002211477 | T0 |
| P3T0 | CASSFGPTYGYTF | 0.003723835 | T0 |
| P2T0 | CASSFRTVSSYEQYF | 0.000369772 | T0 |
| P11T0 | CASSFYPKEKLFF | 5.54E-05 | T0 |
| P3T0 | CASSGEAFYEQYF | 0.005896185 | T0 |
| P10T0 | CASSGRQGAGGELFF | 0.00965364 | T0 |
| P11T0 | CASSHDRSQTQYF | 0.001616766 | T0 |
| P2T0 | CASSHPGTSGRTYTDTQYF | 0.000884352 | T0 |
| P11T0 | CASSHRTGISYEQYF | 0.002022451 | T0 |
| P3T0 | CASSHRTSGSGDTQYF | 0.002036689 | T0 |
| P3T0 | CASSLARGQPQHF | 0.00114304 | T0 |
| P3T0 | CASSLGLKDEQYF | 0.006613444 | T0 |
| P11T0 | CASSLIYQEGRSNTGELFF | 0.001164129 | T0 |
| P12T0 | CASSLLPGGQGVPLHF | 0.016038926 | T0 |
| P1T0 | CASSLQGASSPLHF | 0.000667048 | T0 |
| P1T0 | CASSLQGEYQPQHF | 0.000779149 | T0 |
| P10T0 | CASSLRKLEDEQYF | 0.004355558 | T0 |
| P2T0 | CASSLSGGSGANVLTF | 0.000724992 | T0 |
| P11T0 | CASSLTSTDTQYF | 0.000549219 | T0 |
| P2T0 | CASSLTTELAGETDTQYF | 0.00182429 | T0 |
| P11T0 | CASSLVGPGADIQYF | 0.001331345 | T0 |
| P10T0 | CASSPGLAGNYEQYF | 0.005904902 | T0 |
| P10T0 | CASSPGRWYEQYF | 0.00303979 | T0 |
| P1T0 | CASSPGTGTYGYTF | 0.001554019 | T0 |
| P3T0 | CASSPKTGTIYGYTF | 0.002674264 | T0 |
| P11T0 | CASSPLRSGGSTKVHSSNQPQHF | 0.002313637 | T0 |
| P10T0 | CASSPRTGAGGELFF | 0.142662206 | T0 |
| P10T0 | CASSPTLASSPQYF | 0.00549096 | T0 |
| P10T0 | CASSQDAGGTGELFF | 0.003802581 | T0 |
| P10T0 | CASSQDRGVGSYEQYF | 0.069028889 | T0 |
| P11T0 | CASSQDRNTQYF | 0.000936987 | T0 |
| P3T0 | CASSQEIQPGELFF | 0.004213948 | T0 |
| P11T0 | CASSQGRPPTGELFF | 0.007836507 | T0 |
| P1T0 | CASSQVEGDESPLHF | 0.000987884 | T0 |
| P10T0 | CASSRGLLTDTQYF | 0.008207938 | T0 |
| P10T0 | CASSRTWGTEAFF | 0.005977832 | T0 |

| | | | |
|---|---|---|---|
| P3T0 | CASSRVTGTPSYGYTF | 0.012130635 | T0 |
| P11T0 | CASSSDSLYEQYF | 0.000518741 | T0 |
| P2T0 | CASSSRGTSNQPQHF | 0.00358715 | T0 |
| P1T0 | CASSSSQGAMDTIYF | 0.001890471 | T0 |
| P1T0 | CASSTGQSDRYNEQFF | 0.002149662 | T0 |
| P3T0 | CASSTRAFMNTEAFF | 0.00497486 | T0 |
| P1T0 | CASSVAWDLIYEQYF | 0.006622206 | T0 |
| P3T0 | CASSVEIGGGRGELFF | 0.014281471 | T0 |
| P10T0 | CASSVNWYNEQFF | 0.009178143 | T0 |
| P3T0 | CASSWTSGDTDTQYF | 0.002278456 | T0 |
| P1T0 | CASSYGFYQPQHF | 0.001175219 | T0 |
| P10T0 | CASSYSDRPSTDTQYF | 0.01155335 | T0 |
| P3T0 | CASSYSGDDTQYF | 0.149256048 | T0 |
| P1T0 | CASSYSPSNQPQHF | 0.000794917 | T0 |
| P11T0 | CASTPGDEQYF | 0.000787481 | T0 |
| P1T0 | CATSDWKPDSTNEKLFF | 0.001129793 | T0 |
| P3T0 | CATSRGEGRDTQYF | 0.005066868 | T0 |
| P11T0 | CATSRVAGETQYF | 0.000566517 | T0 |
| P2T0 | CATSSSLAGESGELFF | 0.001204146 | T0 |
| P10T0 | CAWSVPGSVVQPQHF | 0.007448054 | T0 |
| P11T0 | CSAGTGDTQYF | 9.00E-05 | T0 |
| P10T0 | CSARGLGLLYEQYF | 0.037362115 | T0 |
| P1T0 | CSARGQGWTGELFF | 0.000796869 | T0 |
| P3T0 | CSARNPREGTGELFF | 0.010821961 | T0 |
| P1T0 | CSARYLPGQVSSNEQFF | 0.001515501 | T0 |
| P3T0 | CSASSLALVQETQYF | 0.001978937 | T0 |
| P3T0 | CSATGTGVNEQFF | 0.000635591 | T0 |
| P11T0 | CSATSGLTDTQYF | 0.001589377 | T0 |
| P2T0 | CSVAEQILNTEAFF | 0.001545887 | T0 |
| P1T0 | CSVEAGDTQYF | 0.00087751 | T0 |
| P11T0 | CSVEDQGQGYNLGNLNYGYTF | 0.000622942 | T0 |
| P1T0 | CSVEDRGSGFEQYF | 0.001197519 | T0 |
| P1T0 | CSVEFGRGRDEKLFF | 0.003575212 | T0 |
| P1T0 | CSVEGDRGRAEKLFF | 0.00126532 | T0 |
| P11T0 | CSVELGNTGELFF | 0.000348642 | T0 |
| P3T0 | CSVGDSGSGGTQYF | 0.001894136 | T0 |
| P11T0 | CSVGVATGAGELFF | 0.000444606 | T0 |
| P1T0 | CSVNTGVSGANVLTF | 0.003053977 | T0 |
| P10T1 | CAISDRIYGYTF | 0.00397505 | T1 |
| P3T1 | CASDSGLADTQYF | 0.004667522 | T1 |
| P11T1 | CASIPEEGRTQYF | 0.001897333 | T1 |
| P1T1 | CASKLGAAGLFF | 0.003424198 | T1 |
| P2T1 | CASMVRYTGELFF | 0.007000495 | T1 |
| P2T1 | CASRGPTKGSSYEQYF | 0.001664436 | T1 |
| P1T1 | CASRGTSGIYEQYF | 0.015920388 | T1 |
| P10T1 | CASRIPGRRTDTQYF | 0.004960698 | T1 |
| P2T1 | CASRLHPGDTQYF | 0.014784062 | T1 |
| P2T1 | CASRPATGGDFYGYTF | 0.00131588 | T1 |
| P10T1 | CASRPRDRYQETQYF | 0.013722938 | T1 |
| P10T1 | CASSAVTGLAGGPYEQYF | 0.002959995 | T1 |
| P11T1 | CASSDGTGFYGYTF | 0.001874152 | T1 |
| P1T1 | CASSDSTSGGTLPQYF | 0.004787787 | T1 |
| P3T1 | CASSEDYGREETQYF | 0.004054044 | T1 |
| P10T1 | CASSEEPGGIEQYF | 0.003030033 | T1 |
| P10T1 | CASSELAGGPETQYF | 0.00661402 | T1 |
| P2T1 | CASSFAAVLSMNTEAFF | 0.001658874 | T1 |

| | | | |
|---|---|---|---|
| P3T1 | CASSFGPTYGYTF | 0.004059836 | T1 |
| P2T1 | CASSFRTVSSYEQYF | 0.002335796 | T1 |
| P11T1 | CASSFYPKEKLFF | 0.001009076 | T1 |
| P3T1 | CASSGEAFYEQYF | 0.032394788 | T1 |
| P10T1 | CASSGRQGAGGELFF | 0.006822668 | T1 |
| P11T1 | CASSHDRSQTQYF | 0.00448903 | T1 |
| P2T1 | CASSHPGTSGRTYTDTQYF | 0.001709048 | T1 |
| P11T1 | CASSHRTGISYEQYF | 0.007917709 | T1 |
| P3T1 | CASSHRTSGSGDTQYF | 0.005392022 | T1 |
| P3T1 | CASSLARGQPQHF | 0.005510944 | T1 |
| P3T1 | CASSLGLKDEQYF | 0.008343386 | T1 |
| P11T1 | CASSLIYQEGRSNTGELFF | 0.001716642 | T1 |
| P12T1 | CASSLLPGGQGVPLHF | 0.001597733 | T1 |
| P1T1 | CASSLQGASSPLHF | 0.001453144 | T1 |
| P1T1 | CASSLQGEYQPQHF | 0.001439102 | T1 |
| P10T1 | CASSLRKLEDEQYF | 0.004400887 | T1 |
| P2T1 | CASSLSGGSGANVLTF | 0.001558043 | T1 |
| P11T1 | CASSLTSTDTQYF | 0.003095391 | T1 |
| P2T1 | CASSLTTELAGETDTQYF | 0.001867669 | T1 |
| P11T1 | CASSLVGPGADIQYF | 0.001940319 | T1 |
| P10T1 | CASSPGLAGNYEQYF | 0.004104509 | T1 |
| P10T1 | CASSPGRWYEQYF | 0.003068956 | T1 |
| P1T1 | CASSPGTGTYGYTF | 0.002304554 | T1 |
| P3T1 | CASSPKTGTIYGYTF | 0.004787584 | T1 |
| P11T1 | CASSPLRSGGSTKVHSSNQPQHF | 0.011731566 | T1 |
| P10T1 | CASSPRTGAGGELFF | 0.114456122 | T1 |
| P10T1 | CASSPTLASSPQYF | 0.002587479 | T1 |
| P10T1 | CASSQDAGGTGELFF | 0.004301565 | T1 |
| P10T1 | CASSQDRGVGSYEQYF | 0.051709199 | T1 |
| P11T1 | CASSQDRNTQYF | 0.003580969 | T1 |
| P3T1 | CASSQEIQPGELFF | 0.011428591 | T1 |
| P11T1 | CASSQGRPPTGELFF | 0.012776258 | T1 |
| P1T1 | CASSQVEGDESPLHF | 0.001296603 | T1 |
| P10T1 | CASSRGLLTDTQYF | 0.004016901 | T1 |
| P10T1 | CASSRTWGTEAFF | 0.003844126 | T1 |
| P3T1 | CASSRVTGTPSYGYTF | 0.008808102 | T1 |
| P11T1 | CASSSDSLYEQYF | 0.001103183 | T1 |
| P2T1 | CASSSRGTSNQPQHF | 0.004753682 | T1 |
| P1T1 | CASSSSQGAMDTIYF | 0.004964102 | T1 |
| P1T1 | CASSTGQSDRYNEQFF | 0.001083822 | T1 |
| P3T1 | CASSTRAFMNTEAFF | 0.017323069 | T1 |
| P1T1 | CASSVAWDLIYEQYF | 0.008146574 | T1 |
| P3T1 | CASSVEIGGGRGELFF | 0.045923247 | T1 |
| P10T1 | CASSVNWYNEQFF | 0.005427411 | T1 |
| P3T1 | CASSWTSGDTDTQYF | 0.01503205 | T1 |
| P1T1 | CASSYGFYQPQHF | 0.001145292 | T1 |
| P10T1 | CASSYSDRPSTDTQYF | 0.00677923 | T1 |
| P3T1 | CASSYSGDDTQYF | 0.332884386 | T1 |
| P1T1 | CASSYSPSNQPQHF | 0.001159191 | T1 |
| P11T1 | CASTPGDEQYF | 0.002323961 | T1 |
| P1T1 | CATSDWKPDSTNEKLFF | 0.001332711 | T1 |
| P3T1 | CATSRGEGRDTQYF | 0.010594999 | T1 |
| P11T1 | CATSRVAGETQYF | 0.001019208 | T1 |
| P2T1 | CATSSSLAGESGELFF | 0.001989295 | T1 |
| P10T1 | CAWSVPGSVVQPQHF | 0.004269719 | T1 |
| P11T1 | CSAGTGDTQYF | 0.000930014 | T1 |

| | | | |
|---|---|---|---|
| P10T1 | CSARGLGLLYEQYF | 0.034509282 | T1 |
| P1T1 | CSARGQGWTGELFF | 0.001204326 | T1 |
| P3T1 | CSARNPREGTGELFF | 0.006142853 | T1 |
| P1T1 | CSARYLPGQVSSNEQFF | 0.002255622 | T1 |
| P3T1 | CSASSLALVQETQYF | 0.004406947 | T1 |
| P3T1 | CSATGTGVNEQFF | 0.006653557 | T1 |
| P11T1 | CSATSGLTDTQYF | 0.003566538 | T1 |
| P2T1 | CSVAEQILNTEAFF | 0.003404071 | T1 |
| P1T1 | CSVEAGDTQYF | 0.001145937 | T1 |
| P11T1 | CSVEDQGQGYNLGNLNYGYTF | 0.000877818 | T1 |
| P1T1 | CSVEDRGSGFEQYF | 0.001628527 | T1 |
| P1T1 | CSVEFGRGRDEKLFF | 0.006226674 | T1 |
| P1T1 | CSVEGDRGRAEKLFF | 0.00199921 | T1 |
| P11T1 | CSVELGNTGELFF | 0.001094893 | T1 |
| P3T1 | CSVGDSGSGGTQYF | 0.006244309 | T1 |
| P11T1 | CSVGVATGAGELFF | 0.000867225 | T1 |
| P1T1 | CSVNTGVSGANVLTF | 0.001284997 | T1 |
| P10T1 | CAISDRIYGYTF | 0.00397505 | T1 |
| P11T1 | CAISDSGSAGEQYF | 0.000214772 | T1 |
| P5T1 | CASKLDRENSPLHF | 0.006727654 | T1 |
| P1T1 | CASKLGAAGLFF | 0.003424198 | T1 |
| P12T1 | CASKSGREGEQYF | 7.17E-05 | T1 |
| P2T1 | CASMVRYTGELFF | 0.007000495 | T1 |
| P10T1 | CASNPQGQETQYF | 0.001776436 | T1 |
| P5T1 | CASNQDPGTGDVTQYF | 0.001484481 | T1 |
| P2T1 | CASRGPTKGSSYEQYF | 0.001664436 | T1 |
| P1T1 | CASRGTSGIYEQYF | 0.015920388 | T1 |
| P10T1 | CASRIPGRRTDTQYF | 0.004960698 | T1 |
| P2T1 | CASRLHPGDTQYF | 0.014784062 | T1 |
| P10T1 | CASRPRDRYQETQYF | 0.013722938 | T1 |
| P12T1 | CASRQGHSYEQYF | 0.000770825 | T1 |
| P10T1 | CASSAVTGLAGGPYEQYF | 0.002959995 | T1 |
| P11T1 | CASSDGTGFYGYTF | 0.001874152 | T1 |
| P1T1 | CASSDSTSGGTLPQYF | 0.004787787 | T1 |
| P10T1 | CASSEEPGGIEQYF | 0.003030033 | T1 |
| P2T1 | CASSEPGPGDNQPQHF | 0.000638354 | T1 |
| P2T1 | CASSFAAVLSMNTEAFF | 0.001658874 | T1 |
| P5T1 | CASSFRLPGRMAIGELFF | 0.089974902 | T1 |
| P2T1 | CASSFRTVSSYEQYF | 0.002335796 | T1 |
| P12T1 | CASSFTGTSVSDTQYF | 0.000192992 | T1 |
| P12T1 | CASSGALAGGMETQYF | 0.000357942 | T1 |
| P10T1 | CASSGRQGAGGELFF | 0.006822668 | T1 |
| P11T1 | CASSHDRSQTQYF | 0.00448903 | T1 |
| P2T1 | CASSHPGTSGRTYTDTQYF | 0.001709048 | T1 |
| P11T1 | CASSHRTGISYEQYF | 0.007917709 | T1 |
| P11T1 | CASSLAFVGDVSYEQYF | 0.000134789 | T1 |
| P10T1 | CASSLAGTYGYTF | 0.002446184 | T1 |
| P11T1 | CASSLIYQEGRSNTGELFF | 0.001716642 | T1 |
| P5T1 | CASSLLGEETQYF | 0.022852301 | T1 |
| P12T1 | CASSLLPGGQGVPLHF | 0.001597733 | T1 |
| P1T1 | CASSLQGASSPLHF | 0.001453144 | T1 |
| P1T1 | CASSLQGEYQPQHF | 0.001439102 | T1 |
| P5T1 | CASSLSGGTGELFF | 0.01488293 | T1 |
| P10T1 | CASSLSIGGTSYEQYF | 0.00245863 | T1 |
| P11T1 | CASSLTSTDTQYF | 0.003095391 | T1 |
| P2T1 | CASSLTTELAGETDTQYF | 0.001867669 | T1 |

| | | | |
|---|---|---|---|
| P11T1 | CASSLVDGTPVYEQFF | 0.000489263 | T1 |
| P11T1 | CASSLVGPGADIQYF | 0.001940319 | T1 |
| P11T1 | CASSPGISLHEQYF | 0.000857861 | T1 |
| P10T1 | CASSPGLAGNYEQYF | 0.004104509 | T1 |
| P1T1 | CASSPGTGTYGYTF | 0.002304554 | T1 |
| P11T1 | CASSPLRSGGSTKVHSSNQPQHF | 0.011731566 | T1 |
| P10T1 | CASSPPLGDTEAFF | 0.002508047 | T1 |
| P10T1 | CASSPRTGAGGELFF | 0.114456122 | T1 |
| P11T1 | CASSPSPAQPQHF | 0.000530559 | T1 |
| P12T1 | CASSPYGTGSENSPLHF | 0.00092499 | T1 |
| P12T1 | CASSQASGTLETQYF | 0.00074494 | T1 |
| P10T1 | CASSQDAGGTGELFF | 0.004301565 | T1 |
| P10T1 | CASSQDRGVGSYEQYF | 0.051709199 | T1 |
| P11T1 | CASSQDRNTQYF | 0.003580969 | T1 |
| P11T1 | CASSQGRPPTGELFF | 0.012776258 | T1 |
| P1T1 | CASSQVEGDESPLHF | 0.001296603 | T1 |
| P10T1 | CASSRGLLTDTQYF | 0.004016901 | T1 |
| P11T1 | CASSSDSLYEQYF | 0.001103183 | T1 |
| P10T1 | CASSSGLAGGRLTDTQYF | 0.002174332 | T1 |
| P1T1 | CASSSITGDYIFSYEQYF | 0.000484166 | T1 |
| P2T1 | CASSSRGTSNQPQHF | 0.004753682 | T1 |
| P1T1 | CASSSSQGAMDTIYF | 0.004964102 | T1 |
| P1T1 | CASSTGQSDRYNEQFF | 0.001083822 | T1 |
| P1T1 | CASSVAWDLIYEQYF | 0.008146574 | T1 |
| P10T1 | CASSVNWYNEQFF | 0.005427411 | T1 |
| P12T1 | CASSVPDRGDTQYF | 0.00052581 | T1 |
| P12T1 | CASSYRDRASQETQYF | 0.000251739 | T1 |
| P1T1 | CASSYSGDPPTYEQYF | 0.000360223 | T1 |
| P2T1 | CASTGTSGSTGELFF | 0.000449749 | T1 |
| P11T1 | CASTPGDEQYF | 0.002323961 | T1 |
| P12T1 | CASTRLAGSTDTQYF | 0.000427475 | T1 |
| P2T1 | CASWGPMAGVFLDTQYF | 0.001315396 | T1 |
| P2T1 | CATSRDTLAGAPTDTQYF | 0.001796942 | T1 |
| P2T1 | CATSSLGYEQYF | 0.001907204 | T1 |
| P2T1 | CATSSSLAGESGELFF | 0.001989295 | T1 |
| P10T1 | CAWSVPGSVVQPQHF | 0.004269719 | T1 |
| P5T1 | CSAMRGGNTGELFF | 0.003514227 | T1 |
| P10T1 | CSARGLGLLYEQYF | 0.034509282 | T1 |
| P1T1 | CSARGQGWTGELFF | 0.001204326 | T1 |
| P1T1 | CSARYLPGQVSSNEQFF | 0.002255622 | T1 |
| P1T1 | CSASLDSATNEKLFF | 0.001073004 | T1 |
| P11T1 | CSATSGLTDTQYF | 0.003566538 | T1 |
| P10T1 | CSATTSGSITDTQYF | 0.000691467 | T1 |
| P1T1 | CSVEAGDTQYF | 0.001145937 | T1 |
| P1T1 | CSVEDRGSGFEQYF | 0.001628527 | T1 |
| P1T1 | CSVEFGRGRDEKLFF | 0.006226674 | T1 |
| P1T1 | CSVEGDRGRAEKLFF | 0.00199921 | T1 |
| P11T1 | CSVGHSSYEQYF | 0.000840513 | T1 |
| P1T1 | CSVNTGVSGANVLTF | 0.001284997 | T1 |
| P12T1 | CTSRDWGGISEL | 0.000150993 | T1 |
| P10T2 | CAISDRIYGYTF | 0.005537567 | T2 |
| P11T2 | CAISDSGSAGEQYF | 0.000435753 | T2 |
| P5T2 | CASKLDRENSPLHF | 0.010666069 | T2 |
| P1T2 | CASKLGAAGLFF | 0.010839894 | T2 |
| P12T2 | CASKSGREGEQYF | 0.001740968 | T2 |
| P2T2 | CASMVRYTGELFF | 0.010882645 | T2 |

| | | | |
|---|---|---|---|
| P10T2 | CASNPQGQETQYF | 0.005327881 | T2 |
| P5T2 | CASNQDPGTGDVTQYF | 0.007540164 | T2 |
| P2T2 | CASRGPTKGSSYEQYF | 0.003786893 | T2 |
| P1T2 | CASRGTSGIYEQYF | 0.040225179 | T2 |
| P10T2 | CASRIPGRRTDTQYF | 0.007148919 | T2 |
| P2T2 | CASRLHPGDTQYF | 0.007685835 | T2 |
| P10T2 | CASRPRDRYQETQYF | 0.006509144 | T2 |
| P12T2 | CASRQGHSYEQYF | 0.004517475 | T2 |
| P10T2 | CASSAVTGLAGGPYEQYF | 0.007267792 | T2 |
| P11T2 | CASSDGTGFYGYTF | 0.00107737 | T2 |
| P1T2 | CASSDSTSGGTLPQYF | 0.010951105 | T2 |
| P10T2 | CASSEEPGGIEQYF | 0.004744653 | T2 |
| P2T2 | CASSEPGPGDNQPQHF | 0.002141153 | T2 |
| P2T2 | CASSFAAVLSMNTEAFF | 0.002096873 | T2 |
| P5T2 | CASSFRLPGRMAIGELFF | 0.17806067 | T2 |
| P2T2 | CASSFRTVSSYEQYF | 0.001943638 | T2 |
| P12T2 | CASSFTGTSVSDTQYF | 0.002049755 | T2 |
| P12T2 | CASSGALAGGMETQYF | 0.001499773 | T2 |
| P10T2 | CASSGRQGAGGELFF | 0.0065114 | T2 |
| P11T2 | CASSHDRSQTQYF | 0.002828711 | T2 |
| P2T2 | CASSHPGTSGRTYTDTQYF | 0.00140474 | T2 |
| P11T2 | CASSHRTGISYEQYF | 0.000399441 | T2 |
| P11T2 | CASSLAFVGDVSYEQYF | 0.000454319 | T2 |
| P10T2 | CASSLAGTYGYTF | 0.006507452 | T2 |
| P11T2 | CASSLIYQEGRSNTGELFF | 0.000392342 | T2 |
| P5T2 | CASSLLGEETQYF | 0.01068798 | T2 |
| P12T2 | CASSLLPGGQGVPLHF | 0.010359243 | T2 |
| P1T2 | CASSLQGASSPLHF | 0.003779191 | T2 |
| P1T2 | CASSLQGEYQPQHF | 0.002717022 | T2 |
| P5T2 | CASSLSGGTGELFF | 0.012211704 | T2 |
| P10T2 | CASSLSIGGTSYEQYF | 0.003868682 | T2 |
| P11T2 | CASSLTSTDTQYF | 0.000641139 | T2 |
| P2T2 | CASSLTTELAGETDTQYF | 0.001830478 | T2 |
| P11T2 | CASSLVDGTPVYEQFF | 0.00037869 | T2 |
| P11T2 | CASSLVGPGADIQYF | 0.000711649 | T2 |
| P11T2 | CASSPGISLHEQYF | 0.000370363 | T2 |
| P10T2 | CASSPGLAGNYEQYF | 0.0029215 | T2 |
| P1T2 | CASSPGTGTYGYTF | 0.003674546 | T2 |
| P11T2 | CASSPLRSGGSTKVHSSNQPQHF | 0.007841855 | T2 |
| P10T2 | CASSPPLGDTEAFF | 0.004725193 | T2 |
| P10T2 | CASSPRTGAGGELFF | 0.226426421 | T2 |
| P11T2 | CASSPSPAQPQHF | 0.000494045 | T2 |
| P12T2 | CASSPYGTGSENSPLHF | 0.001859282 | T2 |
| P12T2 | CASSQASGTLETQYF | 0.006441185 | T2 |
| P10T2 | CASSQDAGGTGELFF | 0.004853797 | T2 |
| P10T2 | CASSQDRGVGSYEQYF | 0.028638402 | T2 |
| P11T2 | CASSQDRNTQYF | 0.001626908 | T2 |
| P11T2 | CASSQGRPPTGELFF | 0.007090276 | T2 |
| P1T2 | CASSQVEGDESPLHF | 0.004565199 | T2 |
| P10T2 | CASSRGLLTDTQYF | 0.004375341 | T2 |
| P11T2 | CASSSDSLYEQYF | 0.000467698 | T2 |
| P10T2 | CASSSGLAGGRLTDTQYF | 0.006584022 | T2 |
| P1T2 | CASSSITGDYIFSYEQYF | 0.004987513 | T2 |
| P2T2 | CASSSRGTSNQPQHF | 0.009010981 | T2 |
| P1T2 | CASSSSQGAMDTIYF | 0.007515849 | T2 |
| P1T2 | CASSTGQSDRYNEQFF | 0.006002269 | T2 |

| | | | |
|---|---|---|---|
| P1T2 | CASSVAWDLIYEQYF | 0.020997519 | T2 |
| P10T2 | CASSVNWYNEQFF | 0.009846068 | T2 |
| P12T2 | CASSVPDRGDTQYF | 0.00390482 | T2 |
| P12T2 | CASSYRDRASQETQYF | 0.002986894 | T2 |
| P1T2 | CASSYSGDPPTYEQYF | 0.003885522 | T2 |
| P2T2 | CASTGTSGSTGELFF | 0.0017712 | T2 |
| P11T2 | CASTPGDEQYF | 0.001040511 | T2 |
| P12T2 | CASTRLAGSTDTQYF | 0.001717306 | T2 |
| P2T2 | CASWGPMAGVFLDTQYF | 0.001625505 | T2 |
| P2T2 | CATSRDTLAGAPTDTQYF | 0.001190481 | T2 |
| P2T2 | CATSSLGYEQYF | 0.003024134 | T2 |
| P2T2 | CATSSSLAGESGELFF | 0.001443465 | T2 |
| P10T2 | CAWSVPGSVVQPQHF | 0.010158975 | T2 |
| P5T2 | CSAMRGGNTGELFF | 0.010923351 | T2 |
| P10T2 | CSARGLGLLYEQYF | 0.163439169 | T2 |
| P1T2 | CSARGQGWTGELFF | 0.002563098 | T2 |
| P1T2 | CSARYLPGQVSSNEQFF | 0.008133957 | T2 |
| P1T2 | CSASLDSATNEKLFF | 0.00303698 | T2 |
| P11T2 | CSATSGLTDTQYF | 0.000486537 | T2 |
| P10T2 | CSATTSGSITDTQYF | 0.002921077 | T2 |
| P1T2 | CSVEAGDTQYF | 0.00340676 | T2 |
| P1T2 | CSVEDRGSGFEQYF | 0.009042261 | T2 |
| P1T2 | CSVEFGRGRDEKLFF | 0.021524959 | T2 |
| P1T2 | CSVEGDRGRAEKLFF | 0.004431577 | T2 |
| P11T2 | CSVGHSSYEQYF | 0.000568104 | T2 |
| P1T2 | CSVNTGVSGANVLTF | 0.006104805 | T2 |
| P12T2 | CTSRDWGGISEL | 0.001428784 | T2 |
| P4T2 | CAISESSWRGAGELFF | 0.006452435 | T2 |
| P4T2 | CASGLVRSGLETQYF | 0.000951682 | T2 |
| P11T2 | CASIPEEGRTQYF | 0.000264087 | T2 |
| P4T2 | CASKLSGTSPYEQYF | 0.002614529 | T2 |
| P4T2 | CASKTSGTSPQPQHF | 0.000516071 | T2 |
| P2T2 | CASMVRYTGELFF | 0.010882645 | T2 |
| P2T2 | CASRGPTKGSSYEQYF | 0.003786893 | T2 |
| P2T2 | CASRLHPGDTQYF | 0.007685835 | T2 |
| P10T2 | CASRPRDRYQETQYF | 0.006509144 | T2 |
| P12T2 | CASRQGHSYEQYF | 0.004517475 | T2 |
| P11T2 | CASRWGGNTEAFF | 0.000117197 | T2 |
| P10T2 | CASSAVTGLAGGPYEQYF | 0.007267792 | T2 |
| P12T2 | CASSEARAQETQYF | 0.00410162 | T2 |
| P10T2 | CASSEEPGGIEQYF | 0.004744653 | T2 |
| P10T2 | CASSELAGGPETQYF | 0.002262548 | T2 |
| P4T2 | CASSELMGSDQPQHF | 0.002147848 | T2 |
| P2T2 | CASSEPGPGDNQPQHF | 0.002141153 | T2 |
| P2T2 | CASSFAAVLSMNTEAFF | 0.002096873 | T2 |
| P2T2 | CASSFRTVSSYEQYF | 0.001943638 | T2 |
| P12T2 | CASSFTGTSVSDTQYF | 0.002049755 | T2 |
| P11T2 | CASSFYPKEKLFF | 0.000278353 | T2 |
| P12T2 | CASSGALAGGMETQYF | 0.001499773 | T2 |
| P4T2 | CASSGPAGQGGDTQYF | 0.00106696 | T2 |
| P12T2 | CASSGPGLVSNQPQHF | 0.001042215 | T2 |
| P10T2 | CASSGRQGAGGELFF | 0.0065114 | T2 |
| P11T2 | CASSHDRSQTQYF | 0.002828711 | T2 |
| P2T2 | CASSHPGTSGRTYTDTQYF | 0.00140474 | T2 |
| P4T2 | CASSIQPPTGTGGAKNIQYF | 0.000493465 | T2 |
| P10T2 | CASSLAGTYGYTF | 0.006507452 | T2 |

| | | | |
|---|---|---|---|
| P4T2 | CASSLFARGKNIQYF | 0.003611529 | T2 |
| P4T2 | CASSLGGDYGYTF | 0.001041676 | T2 |
| P4T2 | CASSLGNSQETQYF | 0.000533748 | T2 |
| P10T2 | CASSLGYTTDTQYF | 0.000334482 | T2 |
| P11T2 | CASSLIYQEGRSNTGELFF | 0.000392342 | T2 |
| P11T2 | CASSLKGVSSYNEQFF | 0.000321286 | T2 |
| P12T2 | CASSLLPGGQGVPLHF | 0.010359243 | T2 |
| P2T2 | CASSLPEGFLSSYNSPLHF | 0.000560483 | T2 |
| P2T2 | CASSLSSMGNEQYF | 0.0204799 | T2 |
| P4T2 | CASSLTGPEQYF | 0.004121922 | T2 |
| P11T2 | CASSLTSTDTQYF | 0.000641139 | T2 |
| P2T2 | CASSLTTELAGETDTQYF | 0.001830478 | T2 |
| P11T2 | CASSLVDGTPVYEQFF | 0.00037869 | T2 |
| P11T2 | CASSLVGPGADIQYF | 0.000711649 | T2 |
| P4T2 | CASSPDRGPTGELFF | 0.021898767 | T2 |
| P10T2 | CASSPGLAGNYEQYF | 0.0029215 | T2 |
| P11T2 | CASSPLRSGGSTKVHSSNQPQHF | 0.007841855 | T2 |
| P10T2 | CASSPPGTQTQYF | 0.001759556 | T2 |
| P12T2 | CASSPRGQGSNTEAFF | 0.001277202 | T2 |
| P10T2 | CASSPRTGAGGELFF | 0.226426421 | T2 |
| P11T2 | CASSPSPAQPQHF | 0.000494045 | T2 |
| P10T2 | CASSPTLASSPQYF | 0.000442075 | T2 |
| P12T2 | CASSPYGTGSENSPLHF | 0.001859282 | T2 |
| P12T2 | CASSQASGTLETQYF | 0.006441185 | T2 |
| P2T2 | CASSQDLAGIWITDTQYF | 0.00196562 | T2 |
| P2T2 | CASSQDPGDTQYF | 0.00214274 | T2 |
| P10T2 | CASSQDRGVGSYEQYF | 0.028638402 | T2 |
| P12T2 | CASSQDRHRGLNTEAFF | 0.001340108 | T2 |
| P11T2 | CASSQDRNTQYF | 0.001626908 | T2 |
| P12T2 | CASSQFASTDTQYF | 0.002119104 | T2 |
| P11T2 | CASSQGGRASGSDTQYF | 0.000306816 | T2 |
| P11T2 | CASSQGRPPTGELFF | 0.007090276 | T2 |
| P12T2 | CASSRGDEQYF | 0.000853967 | T2 |
| P10T2 | CASSRGLLTDTQYF | 0.004375341 | T2 |
| P12T2 | CASSRQAPPDTQYF | 0.003579516 | T2 |
| P10T2 | CASSRTWGTEAFF | 0.001347799 | T2 |
| P10T2 | CASSSGLAGGRLTDTQYF | 0.006584022 | T2 |
| P11T2 | CASSSGRTDRYF | 0.000288318 | T2 |
| P2T2 | CASSSRGTSNQPQHF | 0.009010981 | T2 |
| P12T2 | CASSSTGTSGTGELFF | 0.002290014 | T2 |
| P4T2 | CASSTSQGAGETQYF | 0.004132849 | T2 |
| P2T2 | CASSVGVIDTDTQYF | 0.000755776 | T2 |
| P10T2 | CASSVNWYNEQFF | 0.009846068 | T2 |
| P12T2 | CASSVPDRGDTQYF | 0.00390482 | T2 |
| P10T2 | CASSWTGYEQYF | 0.001693139 | T2 |
| P12T2 | CASSYRDRASQETQYF | 0.002986894 | T2 |
| P10T2 | CASSYSDRPSTDTQYF | 0.000692654 | T2 |
| P2T2 | CASTGTSGSTGELFF | 0.0017712 | T2 |
| P11T2 | CASTPGDEQYF | 0.001040511 | T2 |
| P4T2 | CASTRGSSGGTEAFF | 0.005219522 | T2 |
| P12T2 | CASTRLAGSTDTQYF | 0.001717306 | T2 |
| P2T2 | CASWGPMAGVFLDTQYF | 0.001625505 | T2 |
| P2T2 | CATSRDTLAGAPTDTQYF | 0.001190481 | T2 |
| P2T2 | CATSRIGGETQYF | 0.001020741 | T2 |
| P11T2 | CATSRVAGETQYF | 0.000327839 | T2 |
| P2T2 | CATSSLGYEQYF | 0.003024134 | T2 |

| | | | |
|---|---|---|---|
| P2T2 | CATSSSLAGESGELFF | 0.001443465 | T2 |
| P10T2 | CAWSVPGSVVQPQHF | 0.010158975 | T2 |
| P4T2 | CSAIGTSGRVGTGELFF | 0.002933042 | T2 |
| P4T2 | CSANLQGARGNQPQHF | 0.001917292 | T2 |
| P10T2 | CSARGLGLLYEQYF | 0.163439169 | T2 |
| P4T2 | CSARGRRDKVTYGYTF | 0.004288731 | T2 |
| P4T2 | CSASQGSISYEQYF | 0.002035355 | T2 |
| P4T2 | CSVDDGVDEQYF | 0.00039715 | T2 |
| P4T2 | CSVDKTGRFNGYTF | 0.001011785 | T2 |
| P12T2 | CSVETGGYTF | 0.001817462 | T2 |
| P11T2 | CSVGGVSPALAKNIQYF | 6.23E-05 | T2 |
| P11T2 | CSVGHSSYEQYF | 0.000568104 | T2 |
| P11T2 | CSVRRGGDTQYF | 0.000144569 | T2 |
| P12T2 | CTSRDWGGISEL | 0.001428784 | T2 |
| P4T3 | CAISESSWRGAGELFF | 0.007396953 | T3 |
| P4T3 | CASGLVRSGLETQYF | 0.003270749 | T3 |
| P11T3 | CASIPEEGRTQYF | 0.001205074 | T3 |
| P4T3 | CASKLSGTSPYEQYF | 0.010141913 | T3 |
| P4T3 | CASKTSGTSPQPQHF | 0.001346067 | T3 |
| P2T3 | CASMVRYTGELFF | 0.023136179 | T3 |
| P2T3 | CASRGPTKGSSYEQYF | 0.013349744 | T3 |
| P2T3 | CASRLHPGDTQYF | 0.008744176 | T3 |
| P10T3 | CASRPRDRYQETQYF | 0.005518441 | T3 |
| P12T3 | CASRQGHSYEQYF | 0.007047698 | T3 |
| P11T3 | CASRWGGNTEAFF | 0.000764641 | T3 |
| P10T3 | CASSAVTGLAGGPYEQYF | 0.010554359 | T3 |
| P12T3 | CASSEARAQETQYF | 0.002207658 | T3 |
| P10T3 | CASSEEPGGIEQYF | 0.010805731 | T3 |
| P10T3 | CASSELAGGPETQYF | 0.006930815 | T3 |
| P4T3 | CASSELMGSDQPQHF | 0.002405914 | T3 |
| P2T3 | CASSEPGPGDNQPQHF | 0.005197049 | T3 |
| P2T3 | CASSFAAVLSMNTEAFF | 0.008802568 | T3 |
| P2T3 | CASSFRTVSSYEQYF | 0.004939034 | T3 |
| P12T3 | CASSFTGTSVSDTQYF | 0.002183343 | T3 |
| P11T3 | CASSFYPKEKLFF | 0.001038077 | T3 |
| P12T3 | CASSGALAGGMETQYF | 0.003499321 | T3 |
| P4T3 | CASSGPAGQGGDTQYF | 0.008522337 | T3 |
| P12T3 | CASSGPGLVSNQPQHF | 0.001423901 | T3 |
| P10T3 | CASSGRQGAGGELFF | 0.015627596 | T3 |
| P11T3 | CASSHDRSQTQYF | 0.005399956 | T3 |
| P2T3 | CASSHPGTSGRTYTDTQYF | 0.003623811 | T3 |
| P4T3 | CASSIQPPTGTGGAKNIQYF | 0.002123518 | T3 |
| P10T3 | CASSLAGTYGYTF | 0.009965995 | T3 |
| P4T3 | CASSLFARGKNIQYF | 0.005460793 | T3 |
| P4T3 | CASSLGGDYGYTF | 0.001925052 | T3 |
| P4T3 | CASSLGNSQETQYF | 0.001627599 | T3 |
| P10T3 | CASSLGYTTDTQYF | 0.004928829 | T3 |
| P11T3 | CASSLIYQEGRSNTGELFF | 0.002293395 | T3 |
| P11T3 | CASSLKGVSSYNEQFF | 0.001084751 | T3 |
| P12T3 | CASSLLPGGQGVPLHF | 0.047966149 | T3 |
| P2T3 | CASSLPEGFLSSYNSPLHF | 0.002840622 | T3 |
| P2T3 | CASSLSSMGNEQYF | 0.035526679 | T3 |
| P4T3 | CASSLTGPEQYF | 0.004641378 | T3 |
| P11T3 | CASSLTSTDTQYF | 0.001289035 | T3 |
| P2T3 | CASSLTTELAGETDTQYF | 0.006673729 | T3 |
| P11T3 | CASSLVDGTPVYEQFF | 0.000903739 | T3 |

| | | | |
|---|---|---|---|
| P11T3 | CASSLVGPGADIQYF | 0.002018637 | T3 |
| P4T3 | CASSPDRGPTGELFF | 0.059655868 | T3 |
| P10T3 | CASSPGLAGNYEQYF | 0.009643481 | T3 |
| P11T3 | CASSPLRSGGSTKVHSSNQPQHF | 0.013945083 | T3 |
| P10T3 | CASSPPGTQTQYF | 0.005809378 | T3 |
| P12T3 | CASSPRGQGSNTEAFF | 0.004796672 | T3 |
| P10T3 | CASSPRTGAGGELFF | 0.141253575 | T3 |
| P11T3 | CASSPSPAQPQHF | 0.000722991 | T3 |
| P10T3 | CASSPTLASSPQYF | 0.006097819 | T3 |
| P12T3 | CASSPYGTGSENSPLHF | 0.004307677 | T3 |
| P12T3 | CASSQASGTLETQYF | 0.009470775 | T3 |
| P2T3 | CASSQDLAGIWITDTQYF | 0.003595143 | T3 |
| P2T3 | CASSQDPGDTQYF | 0.004351422 | T3 |
| P10T3 | CASSQDRGVGSYEQYF | 0.05759416 | T3 |
| P12T3 | CASSQDRHRGLNTEAFF | 0.001928395 | T3 |
| P11T3 | CASSQDRNTQYF | 0.003060152 | T3 |
| P12T3 | CASSQFASTDTQYF | 0.002543513 | T3 |
| P11T3 | CASSQGGRASGSDTQYF | 0.00089964 | T3 |
| P11T3 | CASSQGRPPTGELFF | 0.010214168 | T3 |
| P12T3 | CASSRGDEQYF | 0.001379395 | T3 |
| P10T3 | CASSRGLLTDTQYF | 0.009066849 | T3 |
| P12T3 | CASSRQAPPDTQYF | 0.004784159 | T3 |
| P10T3 | CASSRTWGTEAFF | 0.008595933 | T3 |
| P10T3 | CASSSGLAGGRLTDTQYF | 0.006953406 | T3 |
| P11T3 | CASSSGRTDRYF | 0.000790953 | T3 |
| P2T3 | CASSSRGTSNQPQHF | 0.050885841 | T3 |
| P12T3 | CASSSTGTSGTGELFF | 0.004492952 | T3 |
| P4T3 | CASSTSQGAGETQYF | 0.00868859 | T3 |
| P2T3 | CASSVGVIDTDTQYF | 0.004492302 | T3 |
| P10T3 | CASSVNWYNEQFF | 0.015373603 | T3 |
| P12T3 | CASSVPDRGDTQYF | 0.003457659 | T3 |
| P10T3 | CASSWTGYEQYF | 0.00955736 | T3 |
| P12T3 | CASSYRDRASQETQYF | 0.00223183 | T3 |
| P10T3 | CASSYSDRPSTDTQYF | 0.012292742 | T3 |
| P2T3 | CASTGTSGSTGELFF | 0.003546072 | T3 |
| P11T3 | CASTPGDEQYF | 0.002526238 | T3 |
| P4T3 | CASTRGSSGGTEAFF | 0.007540989 | T3 |
| P12T3 | CASTRLAGSTDTQYF | 0.002080112 | T3 |
| P2T3 | CASWGPMAGVFLDTQYF | 0.002552356 | T3 |
| P2T3 | CATSRDTLAGAPTDTQYF | 0.002894441 | T3 |
| P2T3 | CATSRIGGETQYF | 0.007556114 | T3 |
| P11T3 | CATSRVAGETQYF | 0.00090321 | T3 |
| P2T3 | CATSSLGYEQYF | 0.014048511 | T3 |
| P2T3 | CATSSSLAGESGELFF | 0.002821451 | T3 |
| P10T3 | CAWSVPGSVVQPQHF | 0.01172909 | T3 |
| P4T3 | CSAIGTSGRVGTGELFF | 0.004649277 | T3 |
| P4T3 | CSANLQGARGNQPQHF | 0.002092539 | T3 |
| P10T3 | CSARGLGLLYEQYF | 0.05762711 | T3 |
| P4T3 | CSARGRRDKVTYGYTF | 0.005426111 | T3 |
| P4T3 | CSASQGSISYEQYF | 0.003773333 | T3 |
| P4T3 | CSVDDGVDEQYF | 0.001587856 | T3 |
| P4T3 | CSVDKTGRFNGYTF | 0.001185739 | T3 |
| P12T3 | CSVETGGYTF | 0.001883462 | T3 |
| P11T3 | CSVGGVSPALAKNIQYF | 0.000935208 | T3 |
| P11T3 | CSVGHSSYEQYF | 0.00129591 | T3 |
| P11T3 | CSVRRGGDTQYF | 0.000695753 | T3 |

| | | | |
|---|---|---|---|
| P12T3 | CTSRDWGGISEL | 0.002113669 | T3 |
| P2T4 | CASMVRYTGELFF | 0.021148318 | T4 |
| P2T4 | CASRGPTKGSSYEQYF | 0.009459597 | T4 |
| P2T4 | CASRLHPGDTQYF | 0.004877376 | T4 |
| P2T4 | CASRQAGDTQYF | 0.001747253 | T4 |
| P7T4 | CASRTGTSDHEQYF | 0.037634143 | T4 |
| P7T4 | CASRWGGSYEQYF | 0.006554757 | T4 |
| P7T4 | CASSADSGVHEQYF | 0.003741392 | T4 |
| P7T4 | CASSEDGYSNQPQHF | 0.00031461 | T4 |
| P2T4 | CASSEPGPGDNQPQHF | 0.007194428 | T4 |
| P2T4 | CASSFAAVLSMNTEAFF | 0.002729093 | T4 |
| P2T4 | CASSFRTVSSYEQYF | 0.003133894 | T4 |
| P2T4 | CASSGTSDTDTQYF | 0.003691175 | T4 |
| P7T4 | CASSLGSSYEQYF | 0.096684441 | T4 |
| P7T4 | CASSLLGTGTTNEKLFF | 0.008356079 | T4 |
| P7T4 | CASSLSLYDWGTHTDTQYF | 0.002691653 | T4 |
| P2T4 | CASSLTTELAGETDTQYF | 0.011509848 | T4 |
| P7T4 | CASSLWSATNEKLFF | 0.011399168 | T4 |
| P7T4 | CASSPGHYEQYF | 0.002687811 | T4 |
| P7T4 | CASSPGTGDETQYF | 0.001241041 | T4 |
| P7T4 | CASSPPRTVNSPLHF | 0.043744242 | T4 |
| P7T4 | CASSPQTSGYLYEQYF | 0.003318191 | T4 |
| P7T4 | CASSQDLLANTDTQYF | 0.011423814 | T4 |
| P7T4 | CASSQERTYYDEQYF | 0.012724733 | T4 |
| P2T4 | CASSRSGSRNTGELFF | 0.000869479 | T4 |
| P2T4 | CASSSRGTSNQPQHF | 0.019782038 | T4 |
| P7T4 | CASSSRLAGGTDTQYF | 0.024949135 | T4 |
| P7T4 | CASSVWSGSNEKLFF | 0.023944269 | T4 |
| P7T4 | CASSYPTGGRYEQYF | 0.173018631 | T4 |
| P2T4 | CASTGTSGSTGELFF | 0.002500147 | T4 |
| P7T4 | CATPTGASGRDTTDTQYF | 0.003788873 | T4 |
| P2T4 | CATSRDTLAGAPTDTQYF | 0.003345043 | T4 |
| P2T4 | CATSRIGGETQYF | 0.006502766 | T4 |
| P2T4 | CATSSLGYEQYF | 0.008809713 | T4 |
| P2T4 | CATSSSLAGESGELFF | 0.002011942 | T4 |
| P7T4 | CSARDRTGRSVEAFF | 0.00426318 | T4 |
| P2T5 | CASMVRYTGELFF | 0.010279179 | T5 |
| P2T5 | CASRGPTKGSSYEQYF | 0.005073318 | T5 |
| P2T5 | CASRLHPGDTQYF | 0.01151894 | T5 |
| P2T5 | CASRQAGDTQYF | 0.002731879 | T5 |
| P7T5 | CASRTGTSDHEQYF | 0.0157008 | T5 |
| P7T5 | CASRWGGSYEQYF | 0.004381927 | T5 |
| P7T5 | CASSADSGVHEQYF | 0.01782365 | T5 |
| P7T5 | CASSEDGYSNQPQHF | 0.003527822 | T5 |
| P2T5 | CASSEPGPGDNQPQHF | 0.002990524 | T5 |
| P2T5 | CASSFAAVLSMNTEAFF | 0.004202579 | T5 |
| P2T5 | CASSFRTVSSYEQYF | 0.005786054 | T5 |
| P2T5 | CASSGTSDTDTQYF | 0.002084591 | T5 |
| P7T5 | CASSLGSSYEQYF | 0.010774324 | T5 |
| P7T5 | CASSLLGTGTTNEKLFF | 0.020747219 | T5 |
| P7T5 | CASSLSLYDWGTHTDTQYF | 0.00584776 | T5 |
| P2T5 | CASSLTTELAGETDTQYF | 0.011659271 | T5 |
| P7T5 | CASSLWSATNEKLFF | 0.014120274 | T5 |
| P7T5 | CASSPGHYEQYF | 0.003874838 | T5 |
| P7T5 | CASSPGTGDETQYF | 0.011782006 | T5 |
| P7T5 | CASSPPRTVNSPLHF | 0.042137841 | T5 |

| | | | |
|---|---|---|---|
| P7T5 | CASSPQTSGYLYEQYF | 0.005072134 | T5 |
| P7T5 | CASSQDLLANTDTQYF | 0.016392519 | T5 |
| P7T5 | CASSQERTYYDEQYF | 0.014519254 | T5 |
| P2T5 | CASSRSGSRNTGELFF | 0.00232826 | T5 |
| P2T5 | CASSSRGTSNQPQHF | 0.019603899 | T5 |
| P7T5 | CASSSRLAGGTDTQYF | 0.031272848 | T5 |
| P7T5 | CASSVWSGSNEKLFF | 0.031593883 | T5 |
| P7T5 | CASSYPTGGRYEQYF | 0.105823976 | T5 |
| P2T5 | CASTGTSGSTGELFF | 0.002390862 | T5 |
| P7T5 | CATPTGASGRDTTDTQYF | 0.004294461 | T5 |
| P2T5 | CATSRDTLAGAPTDTQYF | 0.001908017 | T5 |
| P2T5 | CATSRIGGETQYF | 0.002605327 | T5 |
| P2T5 | CATSSLGYEQYF | 0.006111495 | T5 |
| P2T5 | CATSSSLAGESGELFF | 0.002838212 | T5 |
| P7T5 | CSARDRTGRSVEAFF | 0.00375276 | T5 |
| P7T5 | CASRTGTSDHEQYF | 0.0157008 | T5 |
| P7T5 | CASRWGGSYEQYF | 0.004381927 | T5 |
| P7T5 | CASSLGEQFF | 0.003109089 | T5 |
| P7T5 | CASSLGSSYEQYF | 0.010774324 | T5 |
| P7T5 | CASSLLGTGTTNEKLFF | 0.020747219 | T5 |
| P7T5 | CASSLWSATNEKLFF | 0.014120274 | T5 |
| P7T5 | CASSPGHYEQYF | 0.003874838 | T5 |
| P7T5 | CASSPGTGDETQYF | 0.011782006 | T5 |
| P7T5 | CASSPPRTVNSPLHF | 0.042137841 | T5 |
| P7T5 | CASSPQTSGYLYEQYF | 0.005072134 | T5 |
| P7T5 | CASSQDLLANTDTQYF | 0.016392519 | T5 |
| P7T5 | CASSQERTYYDEQYF | 0.014519254 | T5 |
| P7T5 | CASSSGTILEQYF | 0.002816972 | T5 |
| P7T5 | CASSSRLAGGTDTQYF | 0.031272848 | T5 |
| P7T5 | CASSVWSGSNEKLFF | 0.031593883 | T5 |
| P7T5 | CASSYPTGGRYEQYF | 0.105823976 | T5 |
| P7T5 | CASTTDNYSNQPQHF | 0.002973574 | T5 |
| P7T5 | CATPTGASGRDTTDTQYF | 0.004294461 | T5 |
| P7T5 | CSAPSQNTGEETQYF | 0.00052266 | T5 |
| P7T5 | CSVETGTSGTVETQYF | 0.00209153 | T5 |
| P7T6 | CASRTGTSDHEQYF | 0.021351297 | T6 |
| P7T6 | CASRWGGSYEQYF | 0.003349643 | T6 |
| P7T6 | CASSLGEQFF | 0.002954265 | T6 |
| P7T6 | CASSLGSSYEQYF | 0.044344116 | T6 |
| P7T6 | CASSLLGTGTTNEKLFF | 0.008779064 | T6 |
| P7T6 | CASSLWSATNEKLFF | 0.009521408 | T6 |
| P7T6 | CASSPGHYEQYF | 0.002971838 | T6 |
| P7T6 | CASSPGTGDETQYF | 0.004676181 | T6 |
| P7T6 | CASSPPRTVNSPLHF | 0.046808903 | T6 |
| P7T6 | CASSPQTSGYLYEQYF | 0.003027015 | T6 |
| P7T6 | CASSQDLLANTDTQYF | 0.00566199 | T6 |
| P7T6 | CASSQERTYYDEQYF | 0.011609707 | T6 |
| P7T6 | CASSSGTILEQYF | 0.005870838 | T6 |
| P7T6 | CASSSRLAGGTDTQYF | 0.009988569 | T6 |
| P7T6 | CASSVWSGSNEKLFF | 0.023781203 | T6 |
| P7T6 | CASSYPTGGRYEQYF | 0.203460701 | T6 |
| P7T6 | CASTTDNYSNQPQHF | 0.004053241 | T6 |
| P7T6 | CATPTGASGRDTTDTQYF | 0.004695598 | T6 |
| P7T6 | CSAPSQNTGEETQYF | 0.003060578 | T6 |
| P7T6 | CSVETGTSGTVETQYF | 0.002876859 | T6 |
| P6T6 | CASEPGNTGELFF | 0.008578286 | T6 |

| | | | |
|---|---|---|---|
| P1T6 | CASKLGAAGLFF | 0.010186657 | T6 |
| P1T6 | CASRGTSGIYEQYF | 0.030082769 | T6 |
| P7T6 | CASRTGTSDHEQYF | 0.021351297 | T6 |
| P7T6 | CASRWGGSYEQYF | 0.003349643 | T6 |
| P7T6 | CASSADSGVHEQYF | 0.001948951 | T6 |
| P7T6 | CASSAPGTGTHTQYF | 0.002846195 | T6 |
| P7T6 | CASSDSTGPTDTQYF | 0.002805515 | T6 |
| P1T6 | CASSDSTSGGTLPQYF | 0.006447104 | T6 |
| P7T6 | CASSLGSSYEQYF | 0.044344116 | T6 |
| P7T6 | CASSLLGTGTTNEKLFF | 0.008779064 | T6 |
| P1T6 | CASSLPGSEATYEQYF | 0.006063771 | T6 |
| P1T6 | CASSLQGASSPLHF | 0.00612245 | T6 |
| P1T6 | CASSLQGEYQPQHF | 0.006269422 | T6 |
| P7T6 | CASSLTSAAGELFF | 0.00147476 | T6 |
| P7T6 | CASSLWSATNEKLFF | 0.009521408 | T6 |
| P7T6 | CASSPGHYEQYF | 0.002971838 | T6 |
| P7T6 | CASSPGTGDETQYF | 0.004676181 | T6 |
| P1T6 | CASSPGTGTYGYTF | 0.005089352 | T6 |
| P7T6 | CASSPPRTVNSPLHF | 0.046808903 | T6 |
| P6T6 | CASSPRAGAGGELFF | 0.00092123 | T6 |
| P6T6 | CASSQAGQITDTQYF | 0.029263319 | T6 |
| P7T6 | CASSQDLLANTDTQYF | 0.00566199 | T6 |
| P7T6 | CASSQERTYYDEQYF | 0.011609707 | T6 |
| P6T6 | CASSSERDRSNQPQHF | 0.019674345 | T6 |
| P7T6 | CASSSGTILEQYF | 0.005870838 | T6 |
| P1T6 | CASSSITGDYIFSYEQYF | 0.00187069 | T6 |
| P1T6 | CASSSPGGPGNTIYF | 0.004019969 | T6 |
| P7T6 | CASSSRLAGGTDTQYF | 0.009988569 | T6 |
| P1T6 | CASSSSQGAMDTIYF | 0.007739962 | T6 |
| P1T6 | CASSTGQSDRYNEQFF | 0.001647491 | T6 |
| P1T6 | CASSVAWDLIYEQYF | 0.017777088 | T6 |
| P7T6 | CASSVWSGSNEKLFF | 0.023781203 | T6 |
| P1T6 | CASSYGFYQPQHF | 0.00578692 | T6 |
| P7T6 | CASSYPTGGRYEQYF | 0.203460701 | T6 |
| P6T6 | CASSYQGNQPQHF | 0.015101886 | T6 |
| P1T6 | CASSYSGDPPTYEQYF | 0.005140537 | T6 |
| P1T6 | CASSYSPSNQPQHF | 0.009089213 | T6 |
| P7T6 | CAWVLNTEAFF | 0.002645255 | T6 |
| P1T6 | CSVEAGDTQYF | 0.001633781 | T6 |
| P1T6 | CSVEDRGSGFEQYF | 0.00625361 | T6 |
| P1T6 | CSVEFGRGRDEKLFF | 0.016807787 | T6 |
| P1T6 | CSVEGDRGRAEKLFF | 0.004225802 | T6 |
| P1T6 | CSVNTGVSGANVLTF | 0.002297804 | T6 |
| P6T7 | CASEPGNTGELFF | 0.005314134 | T7 |
| P1T7 | CASKLGAAGLFF | 0.00826255 | T7 |
| P1T7 | CASRGTSGIYEQYF | 0.023465608 | T7 |
| P7T7 | CASRTGTSDHEQYF | 0.019874512 | T7 |
| P7T7 | CASRWGGSYEQYF | 0.004944024 | T7 |
| P7T7 | CASSADSGVHEQYF | 0.006304482 | T7 |
| P7T7 | CASSAPGTGTHTQYF | 0.003600325 | T7 |
| P7T7 | CASSDSTGPTDTQYF | 0.003560488 | T7 |
| P1T7 | CASSDSTSGGTLPQYF | 0.005355046 | T7 |
| P7T7 | CASSLGSSYEQYF | 0.036623132 | T7 |
| P7T7 | CASSLLGTGTTNEKLFF | 0.008805342 | T7 |
| P1T7 | CASSLPGSEATYEQYF | 0.003777986 | T7 |
| P1T7 | CASSLQGASSPLHF | 0.002942076 | T7 |

| | | | |
|---|---|---|---|
| P1T7 | CASSLQGEYQPQHF | 0.00364948 | T7 |
| P7T7 | CASSLTSAAGELFF | 0.002836601 | T7 |
| P7T7 | CASSLWSATNEKLFF | 0.009672698 | T7 |
| P7T7 | CASSPGHYEQYF | 0.00325738 | T7 |
| P7T7 | CASSPGTGDETQYF | 0.004565948 | T7 |
| P1T7 | CASSPGTGTYGYTF | 0.004549575 | T7 |
| P7T7 | CASSPPRTVNSPLHF | 0.025663916 | T7 |
| P6T7 | CASSPRAGAGGELFF | 0.005291885 | T7 |
| P6T7 | CASSQAGQITDTQYF | 0.012329853 | T7 |
| P7T7 | CASSQDLLANTDTQYF | 0.011521994 | T7 |
| P7T7 | CASSQERTYYDEQYF | 0.012234709 | T7 |
| P6T7 | CASSSERDRSNQPQHF | 0.006605131 | T7 |
| P7T7 | CASSSGTILEQYF | 0.003902036 | T7 |
| P1T7 | CASSSITGDYIFSYEQYF | 0.004840125 | T7 |
| P1T7 | CASSSPGGPGNTIYF | 0.004250002 | T7 |
| P7T7 | CASSSRLAGGTDTQYF | 0.026099394 | T7 |
| P1T7 | CASSSSQGAMDTIYF | 0.005430111 | T7 |
| P1T7 | CASSTGQSDRYNEQFF | 0.00291935 | T7 |
| P1T7 | CASSVAWDLIYEQYF | 0.013212795 | T7 |
| P7T7 | CASSVWSGSNEKLFF | 0.018700814 | T7 |
| P1T7 | CASSYGFYQPQHF | 0.005079783 | T7 |
| P7T7 | CASSYPTGGRYEQYF | 0.100752419 | T7 |
| P6T7 | CASSYQGNQPQHF | 0.014238609 | T7 |
| P1T7 | CASSYSGDPPTYEQYF | 0.004419071 | T7 |
| P1T7 | CASSYSPSNQPQHF | 0.006935892 | T7 |
| P7T7 | CAWVLNTEAFF | 0.004056677 | T7 |
| P1T7 | CSVEAGDTQYF | 0.004230926 | T7 |
| P1T7 | CSVEDRGSGFEQYF | 0.004065094 | T7 |
| P1T7 | CSVEFGRGRDEKLFF | 0.015330186 | T7 |
| P1T7 | CSVEGDRGRAEKLFF | 0.004133135 | T7 |
| P1T7 | CSVNTGVSGANVLTF | 0.003410029 | T7 |
| P1T7 | CASKLGAAGLFF | 0.00826255 | T7 |
| P1T7 | CASRGTSGIYEQYF | 0.023465608 | T7 |
| P1T7 | CASRPWGTGANTGELFF | 0.001817199 | T7 |
| P1T7 | CASSAGSTDTQYF | 0.000937489 | T7 |
| P1T7 | CASSDSTSGGTLPQYF | 0.005355046 | T7 |
| P1T7 | CASSFVPGQADDEQYF | 0.000181259 | T7 |
| P1T7 | CASSLGEWNEKLFF | 0.002111538 | T7 |
| P1T7 | CASSQVEGDESPLHF | 0.001852184 | T7 |
| P1T7 | CASSRQGANYGYTF | 0.001072745 | T7 |
| P1T7 | CASSSITGDYIFSYEQYF | 0.004840125 | T7 |
| P1T7 | CASSTGQSDRYNEQFF | 0.00291935 | T7 |
| P1T7 | CASSVAWDLIYEQYF | 0.013212795 | T7 |
| P1T7 | CASSYGFYQPQHF | 0.005079783 | T7 |
| P1T7 | CASSYSGDPPTYEQYF | 0.004419071 | T7 |
| P1T7 | CASSYSPSNQPQHF | 0.006935892 | T7 |
| P1T7 | CSVEAGDTQYF | 0.004230926 | T7 |
| P1T7 | CSVEFGRGRDEKLFF | 0.015330186 | T7 |
| P1T7 | CSVEGDRGRAEKLFF | 0.004133135 | T7 |
| P1T7 | CSVNTGVSGANVLTF | 0.003410029 | T7 |
| P1T7 | CSVQDRKNTEAFF | 0.002684582 | T7 |
| P1T8 | CASKLGAAGLFF | 0.002297532 | T8 |
| P1T8 | CASRGTSGIYEQYF | 0.009991918 | T8 |
| P1T8 | CASRPWGTGANTGELFF | 0.001213388 | T8 |
| P1T8 | CASSAGSTDTQYF | 0.001233834 | T8 |
| P1T8 | CASSDSTSGGTLPQYF | 0.004599577 | T8 |

| | | | |
|---|---|---|---|
| P1T8 | CASSFVPGQADDEQYF | 0.001589494 | T8 |
| P1T8 | CASSLGEWNEKLFF | 0.001044961 | T8 |
| P1T8 | CASSQVEGDESPLHF | 0.001083255 | T8 |
| P1T8 | CASSRQGANYGYTF | 0.001520769 | T8 |
| P1T8 | CASSSITGDYIFSYEQYF | 0.001762777 | T8 |
| P1T8 | CASSTGQSDRYNEQFF | 0.002684853 | T8 |
| P1T8 | CASSVAWDLIYEQYF | 0.0063363 | T8 |
| P1T8 | CASSYGFYQPQHF | 0.004281596 | T8 |
| P1T8 | CASSYSGDPPTYEQYF | 0.002639584 | T8 |
| P1T8 | CASSYSPSNQPQHF | 0.001080451 | T8 |
| P1T8 | CSVEAGDTQYF | 0.003290795 | T8 |
| P1T8 | CSVEFGRGRDEKLFF | 0.004263338 | T8 |
| P1T8 | CSVEGDRGRAEKLFF | 0.001803807 | T8 |
| P1T8 | CSVNTGVSGANVLTF | 0.002757886 | T8 |
| P1T8 | CSVQDRKNTEAFF | 0.001305705 | T8 |
