## Supplemental Table 4 for "Low-dose chemotherapy combined with delayed immunotherapy in the neoadjuvant treatment of non-small cell lung cancer and dynamic monitoring of the drug response in peripheral blood"

**Table S4. TCR characteristics across the whole modified therapy**

| Patient | Sample | Shannon | Evenning | Clonity | Time point |
|---|---|---|---|---|---|
| P1 | P1T0 | 9.670422518 | 0.940912842 | 0.059087158 | T0 |
| P1 | P1T1 | 10.28329192 | 0.93021166 | 0.06978834 | T1 |
| P1 | P1T2 | 8.120860561 | 0.870139738 | 0.129860262 | T2 |
| P1 | P1T6 | 9.581860541 | 0.864472995 | 0.135527005 | T6 |
| P1 | P1T7 | 8.587442288 | 0.89680384 | 0.10319616 | T7 |
| P1 | P1T8 | 9.015654215 | 0.943358556 | 0.056641444 | T8 |
| P10 | P10T0 | 6.278333445 | 0.71772799 | 0.28227201 | T0 |
| P10 | P10T1 | 7.298691922 | 0.790284834 | 0.209715166 | T1 |
| P10 | P10T2 | 5.011752146 | 0.628307177 | 0.371692823 | T2 |
| P10 | P10T3 | 6.16781558 | 0.699949468 | 0.300050532 | T3 |
| P11 | P11T0 | 9.139264504 | 0.954383334 | 0.045616666 | T0 |
| P11 | P11T1 | 8.50469813 | 0.938134154 | 0.061865846 | T1 |
| P11 | P11T2 | 10.16238653 | 0.951734488 | 0.048265512 | T2 |
| P11 | P11T3 | 10.26709535 | 0.934149049 | 0.065850951 | T3 |
| P12 | P12T0 | 8.856902032 | 0.938513778 | 0.061486222 | T0 |
| P12 | P12T1 | 8.058678657 | 0.92048776 | 0.07951224 | T1 |
| P12 | P12T2 | 9.666100007 | 0.919646569 | 0.080353431 | T2 |
| P12 | P12T3 | 7.952547259 | 0.913560891 | 0.086439109 | T3 |
| P2 | P2T0 | 7.415680718 | 0.946172795 | 0.053827205 | T0 |
| P2 | P2T1 | 8.26264281 | 0.942318458 | 0.057681542 | T1 |
| P2 | P2T2 | 8.618379051 | 0.931364685 | 0.068635315 | T2 |
| P2 | P2T3 | 7.860103473 | 0.864592628 | 0.135407372 | T3 |
| P2 | P2T4 | 6.959792316 | 0.929701229 | 0.070298771 | T4 |
| P2 | P2T5 | 8.534810194 | 0.917893294 | 0.082106706 | T5 |
| P2 | P2T7 | 8.570130382 | 0.891931043 | 0.108068957 | T7 |
| P3 | P3T0 | 7.497109684 | 0.802480419 | 0.197519581 | T0 |
| P3 | P3T1 | 4.008891897 | 0.630714914 | 0.369285086 | T1 |
| P3 | P3T3 | 6.459351643 | 0.819591716 | 0.180408284 | T3 |
| P3 | P3T5 | 5.786280167 | 0.86258313 | 0.13741687 | T5 |
| P4 | P4T0 | 8.054871931 | 0.932455099 | 0.067544901 | T0 |
| P4 | P4T2 | 8.878353032 | 0.940517096 | 0.059482904 | T2 |
| P4 | P4T3 | 9.36366288 | 0.884860395 | 0.115139605 | T3 |
| P4 | P4T5 | 7.237199332 | 0.947321754 | 0.052678246 | T5 |
| P5 | P5T1 | 8.03527459 | 0.855872367 | 0.144127633 | T1 |
| P5 | P5T2 | 4.536617086 | 0.822830455 | 0.177169545 | T2 |
| P5 | P5T6 | 5.502803253 | 0.834811229 | 0.165188771 | T6 |
| P6 | P6T0 | 5.833181132 | 0.915250031 | 0.084749969 | T0 |
| P6 | P6T2 | 4.308140766 | 0.788483416 | 0.211516584 | T2 |
| P6 | P6T4 | 8.239396678 | 0.867501478 | 0.132498522 | T4 |
| P6 | P6T6 | 4.070208785 | 0.837523848 | 0.162476152 | T6 |
| P6 | P6T7 | 8.753310588 | 0.832194363 | 0.167805637 | T7 |
| P7 | P7T0 | 6.58446061 | 0.858892512 | 0.141107488 | T0 |
| P7 | P7T2 | 8.977450078 | 0.879586823 | 0.120413177 | T2 |
| P7 | P7T4 | 5.769419656 | 0.674174109 | 0.325825891 | T4 |
| P7 | P7T5 | 6.231223762 | 0.77799242 | 0.22200758 | T5 |
| P7 | P7T6 | 5.682305888 | 0.729138812 | 0.270861188 | T6 |
| P7 | P7T7 | 7.001198989 | 0.803163607 | 0.196836393 | T7 |
| P8 | P8T0 | 5.874697466 | 0.853070569 | 0.146929431 | T0 |
| P9 | P9T0 | 8.828587438 | 0.898709066 | 0.101290934 | T0 |
