## Supplemental Table 5 for "Low-dose chemotherapy combined with delayed immunotherapy in the neoadjuvant treatment of non-small cell lung cancer and dynamic monitoring of the drug response in peripheral blood"

Table S5 T cells abundance estimated by [illegible]

| Patient | Stage | Cell types | Value |
|---|---|---|---|
| P5 | c1d0 | CD4+ mer | 6.55E-22 |
| P5 | O1 | CD4+ mer | 0 |
| P4 | c1d0 | CD4+ mer | 3.22E-20 |
| P6 | O1 | CD4+ mer | 2.25E-20 |
| P2 | c1d0 | CD4+ mer | 0 |
| P2 | O1 | CD4+ mer | 0.109364 |
| P7 | c1d0 | CD4+ mer | 2.03E-20 |
| P1 | c1d0 | CD4+ mer | 0.004976 |
| P5 | c1d0 | CD4+ T-ce | 2.02E-19 |
| P5 | O1 | CD4+ T-ce | 3.72E-19 |
| P4 | c1d0 | CD4+ T-ce | 0 |
| P6 | O1 | CD4+ T-ce | 0 |
| P2 | c1d0 | CD4+ T-ce | 4.33E-20 |
| P2 | O1 | CD4+ T-ce | 0.04376 |
| P7 | c1d0 | CD4+ T-ce | 1.87E-19 |
| P1 | c1d0 | CD4+ T-ce | 6.71E-18 |
| P5 | c1d0 | CD4+ naiv | 0.050769 |
| P5 | O1 | CD4+ naiv | 0.021595 |
| P4 | c1d0 | CD4+ naiv | 0 |
| P6 | O1 | CD4+ naiv | 2.84E-19 |
| P2 | c1d0 | CD4+ naiv | 0 |
| P2 | O1 | CD4+ naiv | 0.200849 |
| P7 | c1d0 | CD4+ naiv | 0 |
| P1 | c1d0 | CD4+ naiv | 0.015025 |
| P5 | c1d0 | CD8+ naiv | 2.42E-19 |
| P5 | O1 | CD8+ naiv | 4.09E-20 |
| P4 | c1d0 | CD8+ naiv | 0.000546 |
| P6 | O1 | CD8+ naiv | 0 |
| P2 | c1d0 | CD8+ naiv | 0.000413 |
| P2 | O1 | CD8+ naiv | 0.002434 |
| P7 | c1d0 | CD8+ naiv | 0 |
| P1 | c1d0 | CD8+ naiv | 0 |
| P5 | c1d0 | CD8+ T-ce | 0.075021 |
| P5 | O1 | CD8+ T-ce | 0.087993 |
| P4 | c1d0 | CD8+ T-ce | 1.85E-18 |
| P6 | O1 | CD8+ T-ce | 0 |
| P2 | c1d0 | CD8+ T-ce | 4.22E-20 |
| P2 | O1 | CD8+ T-ce | 0.368443 |
| P7 | c1d0 | CD8+ T-ce | 0 |
| P1 | c1d0 | CD8+ T-ce | 0.01415 |
| P5 | c1d0 | CD8+ Tcm | 0.115713 |
| P5 | O1 | CD8+ Tcm | 0.145014 |
| P4 | c1d0 | CD8+ Tcm | 5.03E-20 |
| P6 | O1 | CD8+ Tcm | 0.046274 |
| P2 | c1d0 | CD8+ Tcm | 0 |
| P2 | O1 | CD8+ Tcm | 0.38659 |
| P7 | c1d0 | CD8+ Tcm | 0 |
| P1 | c1d0 | CD8+ Tcm | 0.072323 |
| P5 | c1d0 | CD8+ Ter | 0.049943 |
| P5 | O1 | CD8+ Ter | 0.046344 |
| P4 | c1d0 | CD8+ Ter | 0 |
| P6 | O1 | CD8+ Ter | 7.09E-18 |
| P2 | c1d0 | CD8+ Ter | 0 |
| P2 | O1 | CD8+ Ter | 0.076355 |
| P7 | c1d0 | CD8+ Ter | 0 |
| P1 | c1d0 | CD8+ Ter | 0.009225 |
