## Supplementary Table 6 for "Low-dose chemotherapy combined with delayed immunotherapy in the neoadjuvant treatment of non-small cell lung cancer and dynamic monitoring of the drug response in peripheral blood"

Table S6 T cells abundance estimated by

| Patient | Stage | Cell types | Value |
|---|---|---|---|
| P5 | c1d0 | CD4_T | 0.15 |
| P5 | O1 | CD4_T | 0.074 |
| P4 | c1d0 | CD4_T | 0.141 |
| P6 | O1 | CD4_T | 0.057 |
| P2 | c1d0 | CD4_T | 0 |
| P2 | O1 | CD4_T | 0.357 |
| P7 | c1d0 | CD4_T | 0.055 |
| P1 | c1d0 | CD4_T | 0.07 |
| P5 | c1d0 | CD8_T | 0.215 |
| P5 | O1 | CD8_T | 0.2 |
| P4 | c1d0 | CD8_T | 0.143 |
| P6 | O1 | CD8_T | 0.093 |
| P2 | c1d0 | CD8_T | 0.076 |
| P2 | O1 | CD8_T | 0.241 |
| P7 | c1d0 | CD8_T | 0 |
| P1 | c1d0 | CD8_T | 0.148 |
| P5 | c1d0 | CD8_naive | 0 |
| P5 | O1 | CD8_naive | 0.001 |
| P4 | c1d0 | CD8_naive | 0.007 |
| P6 | O1 | CD8_naive | 0.003 |
| P2 | c1d0 | CD8_naive | 0.007 |
| P2 | O1 | CD8_naive | 0.001 |
| P7 | c1d0 | CD8_naive | 0 |
| P1 | c1d0 | CD8_naive | 0 |
| P5 | c1d0 | Cytotoxic | 0.01 |
| P5 | O1 | Cytotoxic | 0.016 |
| P4 | c1d0 | Cytotoxic | 0.01 |
| P6 | O1 | Cytotoxic | 0.008 |
| P2 | c1d0 | Cytotoxic | 0 |
| P2 | O1 | Cytotoxic | 0.013 |
| P7 | c1d0 | Cytotoxic | 0 |
| P1 | c1d0 | Cytotoxic | 0.011 |
| P5 | c1d0 | CD4_naive | 0.004 |
| P5 | O1 | CD4_naive | 0.003 |
| P4 | c1d0 | CD4_naive | 0 |
| P6 | O1 | CD4_naive | 0.001 |
| P2 | c1d0 | CD4_naive | 0 |
| P2 | O1 | CD4_naive | 0.016 |
| P7 | c1d0 | CD4_naive | 0 |
| P1 | c1d0 | CD4_naive | 0.001 |
| P5 | c1d0 | CD8+ Tem | 0.003 |
| P5 | O1 | CD8+ Tem | 0.006 |
| P4 | c1d0 | CD8+ Tem | 0.007 |
| P6 | O1 | CD8+ Tem | 0.004 |
| P2 | c1d0 | CD8+ Tem | 0 |
| P2 | O1 | CD8+ Tem | 0.022 |
| P7 | c1d0 | CD8+ Tem | 0 |
| P1 | c1d0 | CD8+ Tem | 0.005 |
