## Supplementary Table 7 for "Low-dose chemotherapy combined with delayed immunotherapy in the neoadjuvant treatment of non-small cell lung cancer and dynamic monitoring of the drug response in peripheral blood"

**Table S7. Annotation of ctDNA somatic mutations.**

| Chr | Pos | Ref | Alt | Impact | Gene |
|---|---|---|---|---|---|
| 9 | 21974721 | C | - | HIGH | CDKN2A |
| 2 | 178098944 | C | T | MODERATE | NFE2L2 |
| 8 | 90955481 | C | T | LOW | NBN |
| 17 | 7577539 | G | A | MODERATE | TP53 |
| 17 | 7578253 | C | A | MODERATE | TP53 |
| 17 | 7577534 | C | A | MODERATE | TP53 |
| 17 | 7578212 | G | A | HIGH | TP53 |
| 17 | 7578253 | C | A | MODERATE | TP53 |
| 17 | 7578271 | T | C | MODERATE | TP53 |
| 2 | 198266834 | T | C | MODERATE | SF3B1 |
| 17 | 7577120 | C | A | MODERATE | TP53 |
| 17 | 7578406 | C | T | MODERATE | TP53 |
| 17 | 7578253 | C | A | MODERATE | TP53 |
| 2 | 202150030 | C | T | HIGH | CASP8 |
| 17 | 7577556 | C | T | MODERATE | TP53 |
| 22 | 50277695 | C | T | HIGH | ZBED4 |
| X | 53225979 | C | T | MODERATE | KDM5C |
| 2 | 202150030 | C | T | HIGH | CASP8 |
| 17 | 7577556 | C | T | MODERATE | TP53 |
| X | 53225979 | C | T | MODERATE | KDM5C |
| 11 | 534289 | C | T | MODERATE | HRAS |
| 3 | 178936091 | G | A | MODERATE | PIK3CA |
| 17 | 7577120 | C | T | MODERATE | TP53 |
| 17 | 7578394 | T | C | MODERATE | TP53 |
| 3 | 178936091 | G | A | MODERATE | PIK3CA |
| 22 | 46932332 | G | A | MODERATE | CELSR1 |
| 1 | 27023141 | GGC | - | MODERATE | ARID1A |
| 3 | 178936091 | G | A | MODERATE | PIK3CA |
| 17 | 7577120 | C | T | MODERATE | TP53 |
| 17 | 7577120 | C | T | MODERATE | TP53 |
| 17 | 7577539 | G | A | MODERATE | TP53 |
| 17 | 7577559 | G | A | MODERATE | TP53 |
| 7 | 98581067 | C | T | HIGH | TRRAP |
| 2 | 209113112 | C | T | MODERATE | IDH1 |
| 9 | 21974792 | G | A | MODERATE | CDKN2A |
| 5 | 112175423 | C | T | HIGH | APC |
| 17 | 7577570 | C | T | MODERATE | TP53 |
| 4 | 1803564 | C | T | MODERATE | FGFR3 |
| 17 | 7577570 | C | T | MODERATE | TP53 |
| 18 | 48591918 | C | T | MODERATE | SMAD4 |
| 17 | 7577570 | C | T | MODERATE | TP53 |
| 17 | 7577570 | C | T | MODERATE | TP53 |
| 17 | 7578190 | T | C | MODERATE | TP53 |
| 17 | 7577570 | C | T | MODERATE | TP53 |
| 17 | 7579358 | C | A | MODERATE | TP53 |
| 1 | 27023874 | A | G | MODERATE | ARID1A |
| 1 | 114968316 | G | A | MODERATE | TRIM33 |
| 5 | 38510808 | G | A | MODERATE | LIFR |
| 9 | 21974792 | G | A | MODERATE | CDKN2A |
| 2 | 25457242 | C | T | MODERATE | DNMT3A |
| 14 | 20825323 | - | [illegible]CACAC[illegible] | MODIFIER | PARP2 |
| 1 | 27023874 | A | G | MODERATE | ARID1A |
| 1 | 51934149 | C | T | MODERATE | EPS15 |
| 17 | 7577545 | T | C | MODERATE | TP53 |

| | | | | | |
|---|---|---|---|---|---|
| 3 | 178916854 | G | A | MODERATE | PIK3CA |
| 1 | 27101099 | C | T | HIGH | ARID1A |
| 2 | 25457243 | G | A | MODERATE | DNMT3A |
| 20 | 57484420 | C | T | MODERATE | GNAS |
| 2 | 25457243 | G | A | MODERATE | DNMT3A |
| 20 | 57484420 | C | T | MODERATE | GNAS |
| 2 | 25457243 | G | A | MODERATE | DNMT3A |
| 20 | 57484420 | C | T | MODERATE | GNAS |
| 1 | 156843455 | T | G | MODERATE | NTRK1 |
| 3 | 128205858 | T | C | MODERATE | GATA2 |
| 5 | 149516594 | G | A | MODERATE | PDGFRB |
| 19 | 50918163 | A | G | MODERATE | POLD1 |
| 17 | 7577094 | G | A | MODERATE | TP53 |
| 1 | 162748481 | G | A | MODERATE | DDR2 |
| 12 | 133263850 | C | T | MODERATE | POLE |
| 17 | 7577559 | G | A | MODERATE | TP53 |
| 17 | 7579377 | G | A | HIGH | TP53 |
| 19 | 1220629 | C | T | MODERATE | STK11 |
| 19 | 13210408 | C | T | MODERATE | LYL1 |
| 20 | 39766334 | C | T | MODERATE | PLCG1 |

| Effect | Transcript | cDNA |
|---|---|---|
| frameshift_variant | NM_000077.4 | c.106del |
| missense_variant | NM_006164.5 | c.101G>A |
| splice_region_variant,synonymous_variant | NM_002485.4 | c.2184G>A |
| missense_variant | NM_000546.5 | c.742C>T |
| missense_variant | NM_000546.5 | c.596G>T |
| missense_variant | NM_000546.5 | c.747G>T |
| stop_gained | NM_000546.5 | c.637C>T |
| missense_variant | NM_000546.5 | c.596G>T |
| missense_variant | NM_000546.5 | c.578A>G |
| missense_variant | NM_012433.3 | c.2098A>G |
| missense_variant | NM_000546.5 | c.818G>T |
| missense_variant | NM_000546.5 | c.524G>A |
| missense_variant | NM_000546.5 | c.596G>T |
| stop_gained | NM_033355.3 | c.1294C>T |
| missense_variant | NM_000546.5 | c.725G>A |
| stop_gained | NM_014838.3 | c.385C>T |
| missense_variant | NM_004187.4 | c.2870G>A |
| stop_gained | NM_033355.3 | c.1294C>T |
| missense_variant | NM_000546.5 | c.725G>A |
| missense_variant | NM_004187.4 | c.2870G>A |
| missense_variant | NM_001130442.2 | c.34G>A |
| missense_variant | NM_006218.4 | c.1633G>A |
| missense_variant | NM_000546.5 | c.818G>A |
| missense_variant | NM_000546.5 | c.536A>G |
| missense_variant | NM_006218.4 | c.1633G>A |
| missense_variant | NM_014246.3 | c.736C>T |
| inframe_deletion | NM_006015.6 | c.258_260del |
| missense_variant | NM_006218.4 | c.1633G>A |
| missense_variant | NM_000546.5 | c.818G>A |
| missense_variant | NM_000546.5 | c.818G>A |
| missense_variant | NM_000546.5 | c.742C>T |
| missense_variant | NM_000546.5 | c.722C>T |
| stop_gained | NM_001244580.1 | c.8986C>T |
| missense_variant | NM_005896.3 | c.395G>A |
| missense_variant | NM_000077.4 | c.35C>T |
| stop_gained | NM_000038.6 | c.4132C>T |
| missense_variant | NM_000546.5 | c.711G>A |
| missense_variant,splice_region_variant | NM_000142.4 | c.742C>T |
| missense_variant | NM_000546.5 | c.711G>A |
| missense_variant | NM_005359.6 | c.1081C>T |
| missense_variant | NM_000546.5 | c.711G>A |
| missense_variant | NM_000546.5 | c.711G>A |
| missense_variant | NM_000546.5 | c.659A>G |
| missense_variant | NM_000546.5 | c.711G>A |
| missense_variant | NM_000546.5 | c.329G>T |
| missense_variant | NM_006015.6 | c.980A>G |
| missense_variant | NM_015906.4 | c.1450C>T |
| missense_variant | NM_001364297.1 | c.749C>T |
| missense_variant | NM_000077.4 | c.35C>T |
| missense_variant | NM_022552.4 | c.2645G>A |
| intron_variant | NM_005484.3 | +15_1467+16insC[illegible] |
| missense_variant | NM_006015.6 | c.980A>G |
| missense_variant | NM_001981.3 | c.305G>A |
| missense_variant | NM_000546.5 | c.736A>G |

| | | |
|---|---|---|
| missense_variant | NM_006218.4 | c.241G>A |
| stop_gained | NM_006015.6 | c.4381C>T |
| missense_variant | NM_022552.4 | c.2644C>T |
| missense_variant | NM_000516.6 | c.601C>T |
| missense_variant | NM_022552.4 | c.2644C>T |
| missense_variant | NM_000516.6 | c.601C>T |
| missense_variant | NM_022552.4 | c.2644C>T |
| missense_variant | NM_000516.6 | c.601C>T |
| missense_variant | NM_002529.3 | c.881T>G |
| missense_variant | NM_032638.5 | c.17A>G |
| missense_variant | NM_002609.4 | c.17C>T |
| missense_variant | NM_002691.4 | c.2480A>G |
| missense_variant | NM_000546.5 | c.844C>T |
| missense_variant | NM_006182.4 | c.2395G>A |
| missense_variant | NM_006231.4 | c.52G>A |
| missense_variant | NM_000546.5 | c.722C>T |
| stop_gained | NM_000546.5 | c.310C>T |
| missense_variant | NM_000455.5 | c.647C>T |
| missense_variant | NM_005583.5 | c.568G>A |
| missense_variant | NM_002660.3 | c.53C>T |

| Protein | source |
|---|---|
| p.Ala36ArgfsTer17 | P1T7 |
| p.Arg34Gln | P1T2 |
| p.Glu728= | P1T2 |
| p.Arg248Trp | P2T0 |
| p.Gly199Val | P2T0 |
| p.Arg249Ser | P2T6 |
| p.Arg213Ter | P2T6 |
| p.Gly199Val | P2T6 |
| p.His193Arg | P2T6 |
| p.Lys700Glu | P2T1 |
| p.Arg273Leu | P2T1 |
| p.Arg175His | P2T1 |
| p.Gly199Val | P2T2 |
| p.Arg432Ter | P3T0 |
| p.Cys242Tyr | P3T0 |
| p.Arg129Ter | P3T0 |
| p.Gly957Asp | P3T0 |
| p.Arg432Ter | P3T1 |
| p.Cys242Tyr | P3T1 |
| p.Gly957Asp | P3T1 |
| p.Gly12Ser | P3T5 |
| p.Glu545Lys | P4T0 |
| p.Arg273His | P4T0 |
| p.His179Arg | P4T7 |
| p.Glu545Lys | P4T1 |
| p.Pro246Ser | P4T1 |
| p.Gly87del | P4T3 |
| p.Glu545Lys | P4T3 |
| p.Arg273His | P4T3 |
| p.Arg273His | P4T5 |
| p.Arg248Trp | P4T5 |
| p.Ser241Phe | P4T5 |
| p.Gln2996Ter | P5T0 |
| p.Arg132His | P5T4 |
| p.Ser12Leu | P5T4 |
| p.Gln1378Ter | P5T6 |
| p.Met237Ile | P6T7 |
| p.Arg248Cys | P6T2 |
| p.Met237Ile | P6T2 |
| p.Arg361Cys | P6T2 |
| p.Met237Ile | P6T4 |
| p.Met237Ile | P6T5 |
| p.Tyr220Cys | P6T5 |
| p.Met237Ile | P6T6 |
| p.Arg110Leu | P6T6 |
| p.Lys327Arg | P7T0 |
| p.Pro484Ser | P7T6 |
| p.Ser250Phe | P7T6 |
| p.Ser12Leu | P7T6 |
| p.Arg882His | P7T1 |
| ACAG | P7T1 |
| p.Lys327Arg | P7T2 |
| p.Arg102Lys | P7T3 |
| p.Met246Val | P7T4 |

| | |
|---|---|
| p.Glu81Lys | P7T5 |
| p.Arg1461Ter | P8T0 |
| p.Arg882Cys | P8T0 |
| p.Arg201Cys | P8T0 |
| p.Arg882Cys | P8T1 |
| p.Arg201Cys | P8T1 |
| p.Arg882Cys | P8T2 |
| p.Arg201Cys | P8T2 |
| p.Val294Gly | P9T0 |
| p.Glu6Gly | P9T0 |
| p.Ala6Val | P9T0 |
| p.Lys827Arg | P9T0 |
| p.Arg282Trp | P9T1 |
| p.Glu799Lys | P9T3 |
| p.Glu18Lys | P9T3 |
| p.Ser241Phe | P9T3 |
| p.Gln104Ter | P9T3 |
| p.Ser216Phe | P9T3 |
| p.Glu190Lys | P9T3 |
| p.Ser18Leu | P9T3 |
